# Breath Volatile Flux Reveals Age-Dependent Metabolic Markers for Breast Cancer Detection

**DOI:** 10.1101/2025.02.12.25322153

**Authors:** Theo Issitt, James Turvill, Jenny Piper, Kelly Redeker

**Affiliations:** Department of Biology, University of York, York YO10 5DD, United Kingdom; Department of Gastroenterology, York Hospital, York Teaching Hospital NHS Foundation Trust, York, UK; Breast Surgery, York Teaching Hospitals NHS Foundation Trust, York, GBR

## Abstract

Breast cancer remains a dominant health risk for women globally with early detection a primary indicator for successful treatment and survival. Current approaches for diagnosis are invasive, costly, and accuracy can be improved. Breath testing using volatile organic compounds (VOCs) as biomarkers presents an exciting avenue for non-invasive cancer diagnosis. However, breath sampling for cancer diagnosis has not yet delivered reliable biomarkers or technology. This is, in-part due to methodology, which introduces confounding factors when attempting to scale to diverse patient populations. We utilize a novel approach to human clinical breath sampling, in which we quantify breath volatile flux. Breath volatile flux considers inhaled air alongside exhaled air to generate a dynamic breath profile which enhances our ability to identify compounds metabolized in humans. We present a novel breath collection platform into which breast cancer clinic patients, in a pilot study of 60 women, provided 5 mins duration breath samples for analysis by gas chromatography mass spectrometry (GC-MS). GC methods targeted a suit of 11 previously identified compounds alongside non-targeted scans. When examining the data across the entire cohort, butanone was the only significantly altered (increased) breath volatile and was able to separate benign tumour and cancer patients from normal patients. However, patient age was observed as a primary confounding factor and reduced the accuracy of butanone’s diagnostic potential. When age was controlled for, chloroform, styrene and isopropyl alcohol acted as indicators of breast cancer health status. Furthermore, once age was accounted for and cancer patients were identified, grade of cancer was indicated by chloroform and DMS fluxes. Using the top 5 discriminate compounds and receiver operator curves we were able to identify cancer from normal patients with area under the curve of 93.4%, grade 2/3 cancers from normal patients with 97.6% AUC, and benign from normal patients with 90.5% AUC. This study suggests that volatile flux measurements from breath allows successful identification, and separation of, cancer, benign tumour and healthy patients.

## Introduction

Breast cancer persists as a predominant health concern for women worldwide, with increasing incidence rates particularly affecting populations in developed nations [1]. The relationship between early detection and patient outcomes is well established, as tumours diagnosed during initial stages demonstrate significantly higher treatment success rates [1, 2].

Contemporary diagnostic approaches rely primarily on mammography and tissue biopsy, methodologies that, while effective, present notable drawbacks. These techniques can cause patient discomfort, require specialized facilities and trained personnel, and incur substantial costs [3, 4]. Furthermore, the invasive nature of these procedures can lead to patient reluctance, potentially delaying crucial early detection [2]. Such limitations have driven the search for alternative diagnostic methods that may complement or potentially replace current practices in specific contexts.

Analysis of volatile organic compounds (VOCs) in human breath has emerged as a promising diagnostic approach [5]. Breath sampling presents a non-invasive, lower cost alternative, potentially reducing both patient anxiety and healthcare costs. The underlying principle affirms that pathological processes, including cancer, alter cellular metabolism in ways that produce distinctive VOC signatures detectable in exhaled breath [5]. This approach has garnered particular interest in breast cancer research, where the development of reliable screening methods remains a priority.

Despite extensive research efforts, breath analysis has yet to yield consistently reliable biomarkers for clinical application [5]. The predominant approach in existing literature involves single-point sampling onto chemical traps (typically sorbent tube such as Tenax) followed by non-targeted analyses. This method typically collects exhaled volatiles over time, generating qualitative or semi-qualitative results [6], which generally do not accurately represent normal physiological breath volatile ranges. Pre-concentration offers the ability to detect very low abundance VOCs, which is valuable in biomarker discovery of exhaled breath VOCs using non-targeting techniques, but makes quantification of results challenging. Without quantification of breath biomarkers, comparisons against what would be normal physiological breath VOC concentrations and deviations from that normal range are difficult to assess. Furthermore, comparisons between cohorts of patients from differing demographics is challenging. Identification of breast cancer-specific markers through the single time-point, untargeted analysis approach is also complicated by significant variability in environmental VOC exposure and individual metabolic differences [5].

In response to these challenges, we present a novel approach to human clinical breath sampling that addresses several key limitations of traditional methods. Our methodology implements concurrent sampling of both inhaled and exhaled breath from breast cancer clinic patients, enabling direct measurement of individual human metabolic flux. We hypothesise that this approach may allow an analysis which addresses changes in background VOC levels and individual metabolic variations (Fig 1). Through this refined methodology, we aim to establish a more robust foundation for breath-based cancer diagnostics and work towards establishing normal physiological breath volatile ranges.

**Figure 1.**
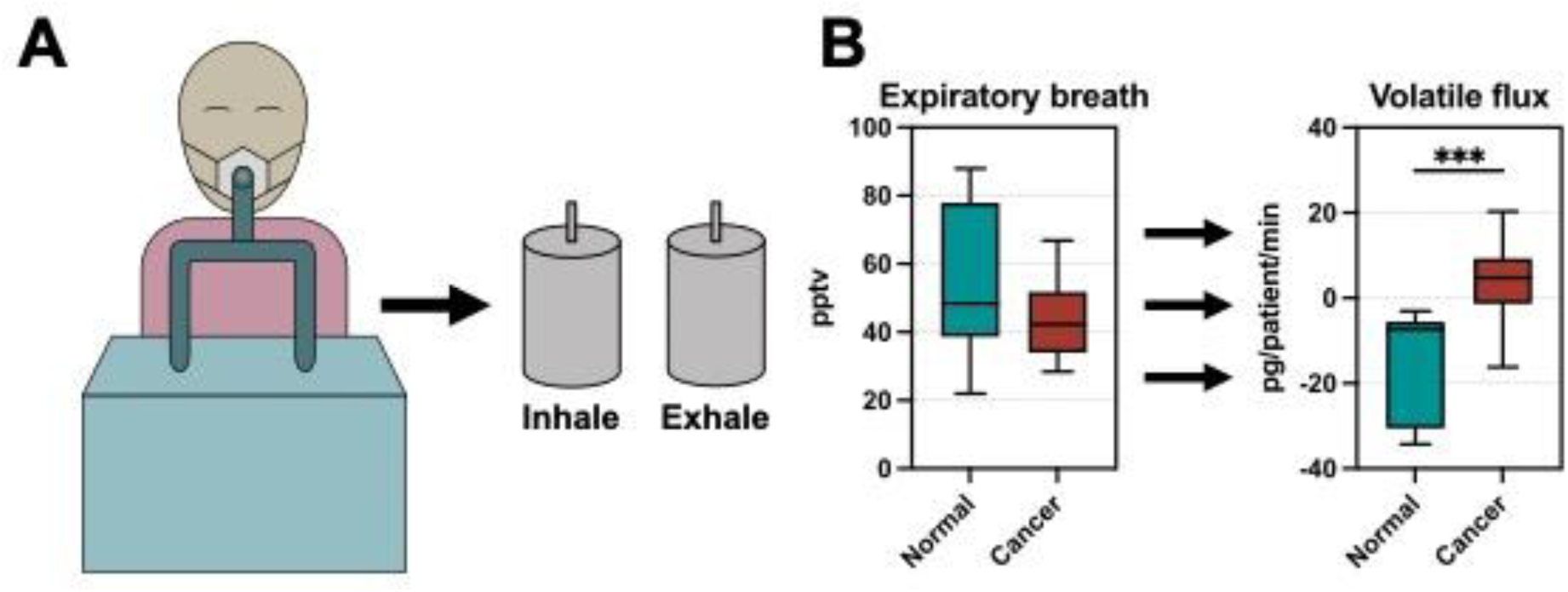
Overview of breath volatile flux. **(A)** Breath Is captured over 5 mins of tidal breathing at rest. Air is drawn into the circuit through a one way valve, driven by patients breathing a T-valve. Two samples are generated, an inhale and exhale sample. **(B)** An example of a single volatile compound (chloroform) in parts per trillion volume in the exhaled breath of normal patients and breast cancer patients with grade 2 or 3 tumours vs the volatile flux of the same compound in pg/patient/min. Unpaired t-test, n=8 for normal patients and n=14 for cancer patients; *** = p<0.001.

Further to our breath sampling approach we present a suite of targeted compounds which we have identified using *in vitro* and *in vivo* models [7, 8] and through a previous review where we identified functional biomarkers of cancer in breath [5]. Taken together, this study aims to characterise metabolic volatile flux in the breath of breast cancer patients in a limited study with a total of 60 women.

## Methods

### Pilot Clinical Study Design and implementation

This observational pilot study was conducted at the Breast Clinic of York and Scarborough Teaching Hospitals NHS Foundation Trust. This study was conducted as a double-blind study in that information beyond patient number were not made available to sample collection staff or sample analysis staff until the completion of the trial.

### Participant Selection

Participants were recruited from the “two-week wait” referral pathway for suspected breast cancer. Eligible participants included adults aged 18 years or older referred for suspected breast cancer. Patients with an existing cancer diagnosis within the past five years (excluding non-melanoma skin cancer), those unable to provide breath samples, and individuals who had eaten within one hour of sampling were excluded. Recruitment aimed to include 20 patients each with breast cancer (“cancer” group), benign breast disease (“benign” group), and no breast disease (“normal” group). Patients who were eligible for inclusion were enrolled into groups based on initial clinician assessment and breath sampling followed, prior to any invasive procedure.

### Patient and Clinical Data Collection

Participants completed a questionnaire with researcher support to record demographic, dietary, exercise, medical, family, and drug history. These data, along with VOC analyses, were reconciled with final clinical diagnoses obtained from routine clinical care.

### Breath Collection and Analysis Procedures

Breath samples were collected using a custom sampling platform (described below) in a quiet, well-ventilated room within the Magnolia Centre at York Hospital. Every patient was sampled in the same room and had generally been in the clinic for at least 20 mins prior. Patients drank water before sampling and had fasted for at least one hour prior. Participants sat at rest for five minutes before breathing normally into a t-valve connected to the breath collection device for an additional five minutes for roughly a 30 minute sampling period per patient, which fit within the usual clinic timeline for patient treatment.

### Breath Sampling Platform

A new breath platform was developed to collect breath from patients at rest for the calculation of breath volatile flux. All materials were stainless steel or silicone except for purchased valves and constructed to fit with standard respiratory circuit equipment (22mm inner diameter). The outline of this device is shown in Fig 1 and a picture of the collection device set up for collection shown in Fig s1. In short, air is drawn into the circuit through a sterile, one-way valve (Intersurgical, UK), then through stainless steel ball valves (RS components, UK) welded to a stainless-steel top plate and mixed in the 5L stainless steel ‘inhale’ reservoir (gastronorm pan, Nisbets, UK). Top plates are screwed onto silicone gaskets prior to sampling, which, with one way valves, creates a single direction, air tight breathing circuit.

Air flows into the reservoirs at the bottom of the tank through a stainless steel tube and exits at the top of the tank to allow even mixing within the chamber. Inhaled air then passes through a silicone tube (Intersurgical, UK) to a single direction T-valve directly into the mouth, allowing inhaled air to be drawn to the lungs through the circuit in one direction. Breath then exits the patient, flows through the T-valve in the other direction, and is directed to the bottom of the 5L stainless steel ‘exhale’ reservoir, which sits in an ice bath and is partially filled with 316 stainless steel ball bearings, increasing surface area for water vapour condensation. Excess exhaled breath is released from the stainless steel “exhalation” reservoir via pressure differential with ambient air. Patients wore a nose clip and breath only through the device during collection. All valves are open except sampling port valves (Swagelok, UK) during sampling (not involved in patient breath dynamics, and shown in Fig s1).

Once the 5min patient breathing is complete all valves are closed and 500mL stainless steel electro-polished evacuated canisters (Lab Commerce, USA) are attached to the sampling ports for inhaled air and exhaled breath sample collection. Samples were transferred into the 500mL canisters within 10 minutes of completion and transferred to the Redeker lab at the University of York for analysis. Samples were stored at 20° C for up to 1 week, a time frame in which the compounds analyzed have been shown to be stable [9].

#### Infection Mitigation Measures

Single-use valves (Intersurgical, UK) or autoclavable materials were employed to minimize transmission risk. Valves were replaced for each participant for new sterile valves, and the collection device was autoclaved between patients.

### Compound selection and identification

This study has two analytical approaches using GC/MS, a targeted compound suite and untargeted discovery. This study presents 11 compounds which were previously identified as modulated in breast cancer models and attempts to follow guidelines of inclusion of a wide range of functional groups [5]. We have previously shown changes in breast cancer cell volatile flux relative to control cells and in response to chemotherapeutic stress [8] and hypoxia [7]. Changes in methyl chloride previously observed under cellular stress [7, 8] made this VOC a target compound along with other chloride containing compounds dichloromethane (DCM) and chloroform. Methyl Iodide, due to previously published links with methyl chloride metabolism in eukaryotes [10], was included as this compound has not been targeted in human breath before and iodine is involved in hormonal homeostasis [11]. Dimethyl sulfide (DMS) has been observed in the breath of patients [5] and acetone is both a potential indicator of glucose metabolism but as it is present ‘on-breath’ in significant concentrations [5], also provided an indication of successful sample collection. 3-methyl pentane and n-hexane have been observed to alter in headspace of breast cancer cells under stress [7, 8], and 2-methyl pentane, while not yet seen to be changed in our previous invitro models, was quantified to investigate if 6 carbon alkane isomers revealed consistent changes. Butanone was observed as a key biomarker in also observed in multiple cancer studies [5]. Styrene was included as we previously identified this compound as released by cells under hypoxic stress [7].

Compound identification was confirmed using purchased standards as previously described [7, 8]. Calibration was performed using standard gases (BOC Specialty Gases, UK). Linear regression of calibration curves confirmed strong, positive linear relationships between observed compound peak areas and moles of gas injected for each VOC (r2 > 0.9 in all cases). For compounds not purchased in gaseous state (BOC Specialty gases, as above), 1-2 mL of compound in liquid phase was injected neat into butyl sealed Wheaton-style glass vials (100 mL) and allowed to equilibrate for 1 h. One mL of headspace air was then removed from neat vial headspace using a gas tight syringe (Trajan, SGE) and injected into the headspace of a second 100 mL butyl sealed Wheaton-style glass vial. This was then repeated, and 1 mL of the 2nd serial dilution vial was injected into the GC/MS system with 29 mL of lab air to give ppb concentrations. This was performed for acetone (Sigma-Aldrich, USA), 2-& 3-methyl pentane, n-hexane (Thermo Scientific, USA, butanone (Sigma-Aldrich, USA) and styrene (Sigma-Aldrich, USA). Reported compounds detected by the GC/MS were confirmed by matching retention times and mass–charge (m/z) ratios with known standards (see table 1).

**Table 1.** Retention times and mass charge ratios used to characterise individual VOCs.

| Compound | Abbreviation | CAS | Retention time | Mass charge ratio ($m/z$) |
| --- | --- | --- | --- | --- |
| Methyl chloride | MeCl | 74-87-3 | 7.8 | 52,50 |
| Trichloroflouromethane | CFC-11 | 75-69-4 | 15.0 | 101.103 |
| Methyl iodide | MeI | 74-88-4 | 15.2 | 127, 142 |
| Dichloromethane | DCM | 75-09-2 | 15.6 | 49, 84 |
| Dimethyl sulfide | DMS | 75-18-3 | 16.2 | 62, 47 |
| Acetone |  | 67-64-1 | 18.2 | 58 |
| Trichloromethane | Chloroform | 67-66-3 | 25.4 | 83, 85 |
| 2- Methyl pentane | 2MP | 107-83-5 | 27.6 | 43, 57 |
| 3- Methyl pentane | 3MP | 96-14-0 | 28 | 43, 57 |
| Butanone |  | 78-93-3 | 28.2 | 57, 72 |
| n-Hexane | nHexane | 110-54-3 | 28.6 | 43, 57 |
| Styrene |  | 100-42-5 | 33.4 | 104, 78 |

### GC-MS method

Our analytical methods have been reported previously [5, 8]. Briefly, air/breath samples were driven from canisters through fine mesh Ascarite® traps [12] to a liquid nitrogen trap, where gases with boiling points greater than ∼-190° C condensed, through pressure differential. Injected volumes were calculated from pressure changes and known condensation trap system volume. Analysis was performed on an Agilent/HP 5972 MSD (Santa Clara, CA, United States) equipped with a PoraBond Q column (25 m × 0.32 mm × 0.5 μm, Restek©, Bellefonte, PN, United States). Both targeted (SIM) and untargeted (SCAN, 45-200 amu) analyses were conducted using electron impact ionization (70 eV), with system temperatures maintained at 250° C (transfer line) and 280° C (ion source). Chromatographic separation employed temperature programming: 35°C (2 min), 10°C/min to 155°C, 1°C/min to 131°C, and 25°C/min to 250°C (5.5 min hold). All samples were analysed within 6 days of collection, with specific SIM parameters and significant SCAN identifications provided in Table 1.

### Data Analysis

Calculation of VOC flux for targeted compounds has previously been described in detail by our group [5, 8]. All targeted compounds have been confirmed and quantified through known standard calibration. All individual sample concentrations have been corrected to CFC-11 concentrations equivalent to the ambient atmospheric concentration of 240 pptv [13, 14].

Untargeted data were classified by retention time and total ion count with 83 compounds identified. As with targeted compounds, untargeted compound fluxes were normalized to CFC-11 as an internal standard. Data matrices were handled in Rstudio v12.1 (Posit Software) where compounds with 50% or above frequency of observation (FOO) of fluxes within any group were included resulting in 46 compounds retained. Missing values were imputed with 1/5^th^ the lowest value, data log transformed (base10) mean-centered and divided by the standard deviation of each variable to transform data in Metaboanalyst following methods previously reported [15]. Extracted data files (.D) were analysed in MzMine 3 [16] for the purpose of compound identification against the Massbank of North America (MoNA) GC-MS library, NIST and Golm libraries were found to have poor match % or produced unlikely compound matches. In MzMine, data were deconvolved and aligned where possible (this was not possible in all files due to file compatibility issues),

Data matrices of flux values were uploaded to Metaboanalyst 6.0 [17] for multivariate analysis. Data were log transformed (base 10) and mean-centred and divided by the standard deviation of each variable. Multivariate analysis was performed in Metaboanalyst using unsupervised principal component analysis (PCA) and supervised partial least-squares discrimination analysis (PLS-DA). PLS-DA was tuned using five-fold cross validation. Variable importance projection (VIP) scores were generated from PLS-DA in Metaboanalyst. Random forest analysis was performed using 2000 feature trees, 7 predictors and random settings. Multivariate exploratory receiver operator curves (ROC) were generated using random forest models and random forest feature selection were used to identify top targets for targeted ROC analysis using selected features and random forest algorithms in Metaboanalyst. Graphpad Prism (GraphPad Software, version 10, San Diego, CA, USA) was used to generate graphs and perform t-tests or ANOVA.

### Ethical Approval

This study received ethical approval from an NHS Research Ethics Committee (IRAS ID: 318636) and University of York Biology Ethics Committee (reference: KR202302). All participants provided written informed consent before any study procedures were undertaken. The study adhered to the ethical principles outlined in the Declaration of Helsinki and Good Clinical Practice guidelines. Approval covered patient recruitment, breath collection, data handling, and analysis as described in the protocol.

## Results

Analysis of breath volatile flux in this patient cohort revealed three key findings. First, the variability in inhaled breath composition significantly impacts the identification of metabolically relevant compounds, with the remaining subset of volatiles with consistent differences between inhaled and exhaled breath not useful for diagnostic purposes. Second, patient age emerged as a critical factor influencing VOC flux outcomes, particularly for chloroform fluxes. Third, after controlling for age, we identified specific volatile biomarkers associated with cancer status.

### Inhaled air vs exhaled breath volatile concentrations

Variability in compound concentration of inhaled breath is substantial, in some cases dwarfing the variability in exhaled breath [18]. As concentration of compounds in the atmosphere varies considerably [13] we aimed to characterize the ‘average’ input of air into the patient’s body over 5 mins; the time in which atmospheric compounds were able to circulate and equilibrate throughout the body. Since blood circulates through the body approximately once every minute, we estimated that 5 minutes would be sufficient to allow tissues in the body in direct contact with blood to equilibrate to the air inhaled by the patients.

We present 11 compounds, arranged by retention time, in Fig 2. While all inhaled air samples were within the normal range of urban air samples for these compounds, as previously reported [13], our analysis revealed particularly high variability (defined as: standard deviation > 50% the mean concentration for any given compound) in ambient (In) concentrations of MeI (Fig 2B), DCM (Fig 2C), 2MP (Fig 2G), 3MP (Fig 2H), butanone (Fig 2I), n-hexane (Fig 2J), and styrene (Fig 2K). In contrast, MeCl (Fig 2A), DMS (Fig 2D), acetone (Fig 2E), and chloroform (Fig 2F) showed relatively low variability in ambient concentrations (Fig 2).

**Figure 2.**
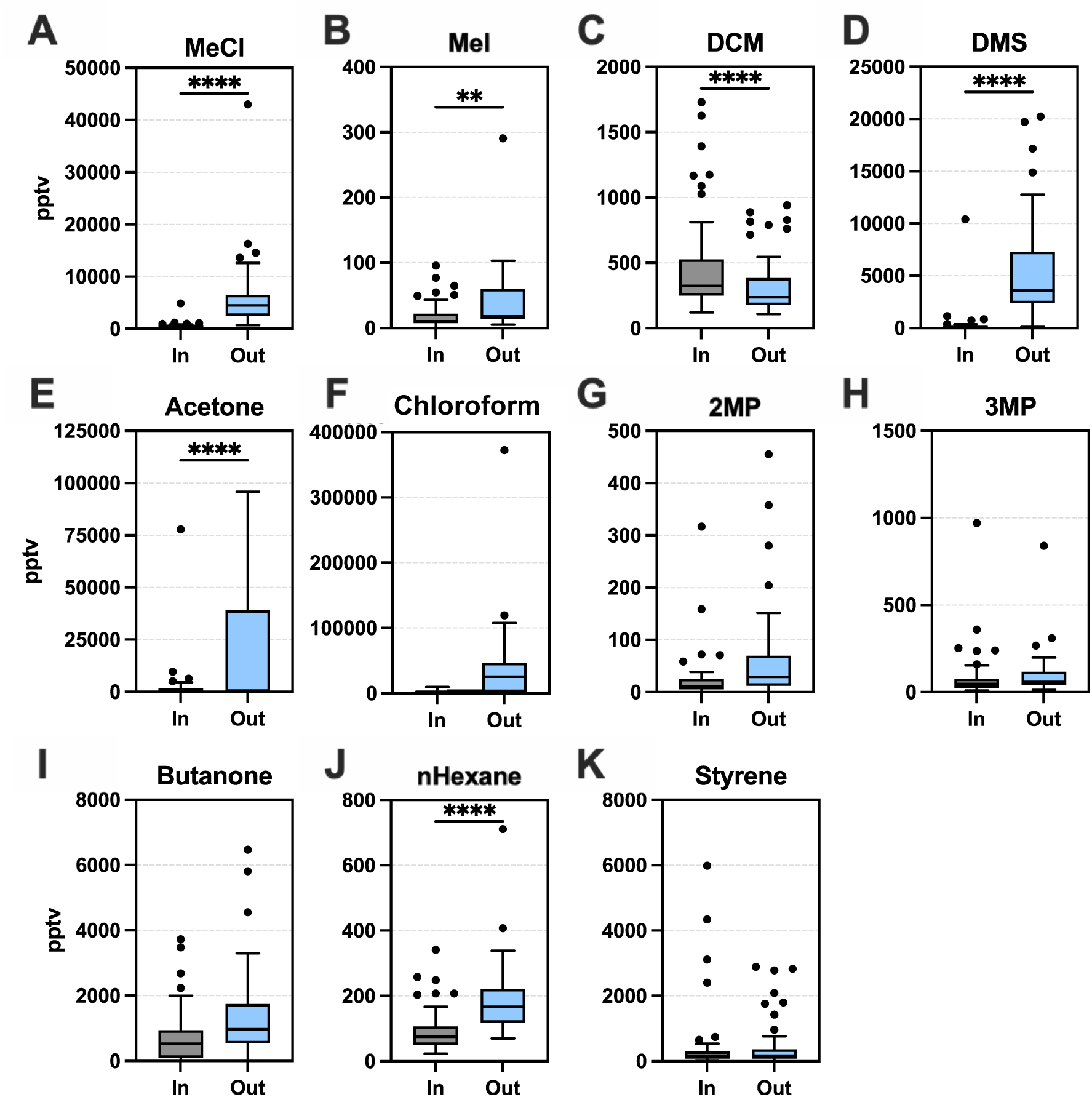
Targeted compound concentrations in Inhaled (In) versus exhaled (Out) breath. Boxplots for 60 female patients, mixed ages (19-94 years). Boxplots show median ± Tukey distribution, in and out breath sample are collected at the same time for each patient. Paired t-test was performed; ****p = <0.0001, **p = 0.01.

This substantial background variability in inhaled air means that only a few compounds demonstrate significant differences between inhaled air and exhaled breath across all patient samples. Key compounds with consistent differences include MeCl (Fig 2A), MeI (Fig 2B), DCM (Fig 2C), DMS (Fig 2D), acetone (Fig 2E), chloroform (Fig 2F), and n-hexane (Fig 2J). Among these, DCM was the only compound with a lower concentration in exhaled breath (Fig 2C). Notably, apart from MeI and DCM, these compounds have been previously identified as metabolically produced by humans, though they have not demonstrated consistent utility in disease diagnosis. This is the first reported instance of MeI being produced through human metabolism and evident in human breath as well as the first description of DCM being utilized in human metabolism or consumed through (bio)chemical processes in the human body.

Of the 12 previously targeted compounds (Figure s2A-K), only 6 were identified as significantly different between inhaled air and exhaled breath in the subset of patients identified as “normal”, MeCl (Fig s2A), MeI (Fig s2B), DCM (Fig s2C), DMS (Fig s2D), acetone (Fig s2E) and n-hexane (Fig s2J). Only DCM was comparatively lower between exhaled breath (out) than inhaled air (Fig s2C). There were no significant changes between inhaled and exhaled samples for chloroform (Fig s2F), 2MP (Fig s2G), 3MP (Fig s2H), butanone (Fig s2I) or styrene (Fig s2K).

Eight of the targeted compounds were significantly different between ‘in’ and ‘out’ breath in patients with cancer. These included MeCl (Fig s3A), MeI (Fig s3B), DCM (Fig s2C), DMS (Fig s2D), acetone (Fig s3E), 2MP (Fig s3G), butanone (Fig s3I) and n-hexane (Fig s3J). In the benign group, 6 compounds were significantly different between ‘in’ and ‘out’ breath, including MeCl (Fig s4A), DMS (Fig s4D), acetone (Fig s4E), 2MP (Fig s4G), butanone (Fig s4I) and n-hexane (Fig s4J).

Six of eight compounds which were statistically different in exhaled breath compared with inhaled air within the cancer group (MeCl, MeI, DCM, DMS, acetone, 2MP, butanone, and n-hexane) were identical to those showing differences in the normal group. Similarly, six of eight compounds in the cancer group overlapped entirely those compounds that were statistically different in the benign group. Compounds which were different in the normal group relative to those identified in benign included MeI, DCM, 2MP and butanone.

### Flux of chloroform is indicative of age

Quantification of metabolite flux within a human body requires measurement of the change in compound concentration between in vs out (exhaled) breath relative to the time spent in the body and an indication of organism or human mass. Units for these metabolic fluxes are generally of the form: mass of compound per biomass per time, or in this paper as µg patient^−1^ min^−1^. Volatile fluxes were calculated for all patients and those compounds targeted for analysis were investigated to determine if they were capable of distinguishing normal patients from patients with benign or cancerous tumours. We discovered that butanone flux was increased in both benign and cancer groups compared with normal (Fig 3A), however no other compounds were shown to be significantly different between groups (Fig s5A-K). Unsupervised principal component analysis (PCA) was performed on targeted compound flux data for all 3 groups; normal, benign and cancerous. No group separation was observed for targeted fluxes using PCA (Fig 3B) or PLS-DA (Fig 3C) and therefore, compounds which show significance when using fluxes across all patients are not explanatory for patient status. Untargeted analysis also revealed no separation by PCA of groups (Fig s6B)

**Figure 3.**
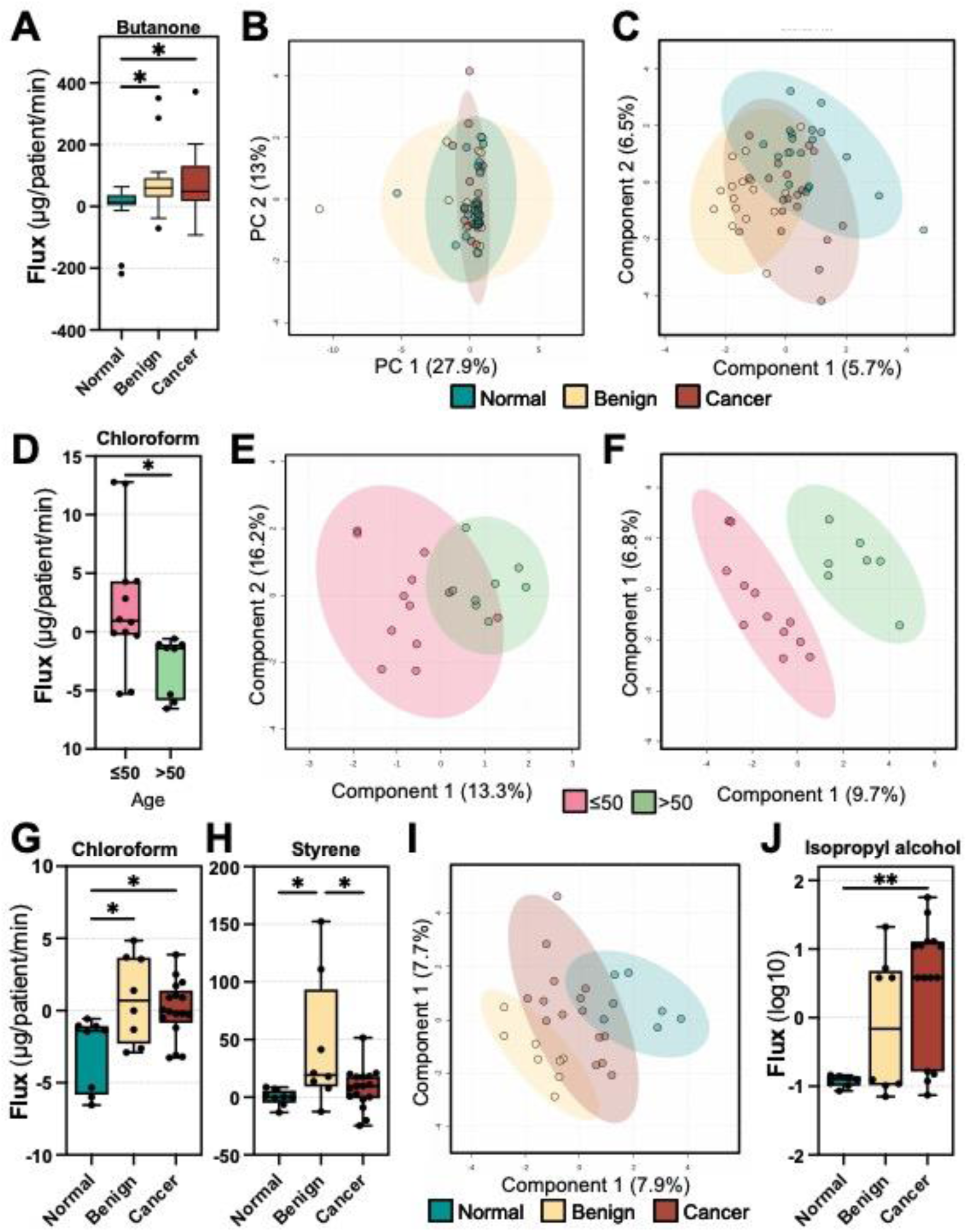
Volatile flux in the breath of normal, benign and breast cancer patients. **(A)** Butanone flux is significantly increased in benign and cancer patients vs normal patients in µg/patient/min. **(B)** Principle component analysis and **(C)** partial least squared discriminant analysis (PLS-DA) of 12 select volatile compound flux values. **(D)** Boxplot of normal group chloroform flux values in µg/patient/min for 50 and under and over 50s. **(E)** PLS-DA for 12 select volatile compounds and **(F)** untargeted analysis for normal 9roup based on age, 50 and under vs over 50 for select volatile flux. **(G)** Chloroform and **(H)** styrene flux, in µg/patient/min in the over 50s. PLS-DA of untargeted analysis in the over 50s. **(J)** lsopropyl alchohol flux(log10) in the over 50s. Boxplots show median ± Tukey distribution (n= 20, 21, 19 for normal, benign and cancer groups respectively). Unpaired t-test was performed for **D**; ** p = < 0.01, *p = <0.05.

In examining volatile metabolite fluxes for all patients, it became apparent that confounding variables contributed to the lack of discriminatory capacity in concentration and flux assessments. In particular, it became clear that age of patient had a substantial impact on individual metabolism. This is supported by our observation of chloroform being increased in the headspace of aging cells and flies [19] and published data showing that metabolism profoundly changes as humans age [20].

Age distribution of patients was not even across groups (Fig s6C). Average (mean) age of all patients was 54, with average age of cancer patients higher than normal or benign (cancer, 66; benign, 50; normal, 49). Age is shown in Fig s6C; considering uncancerous controls, we split the patient population into two groups-age 50 or greater (>50) or those aged 50 or less 50 (≤50).

Targeted compound volatile flux profiles for the normal patients (>50 versus ≤50) were separable using PLS-DA (Fig 3E) but not by PCA (Fig s7A). Separation of these groups was driven by chloroform flux, the only significant change, where ≤50 showed production (on average) while those >50 showed consumption of chloroform (Fig 3D). In patients separated by age, benign and cancer patient populations did not exhibit significant differences in chloroform flux (Fig s7B and C respectively).

In the normal group, age split corresponded with menopause status (table 1). Menopause status in the benign and cancer groups was not split at age 50 however (table 1), so patients were also separated on menopause status, to determine if chloroform changes were menopause dependent. Only the normal group showed significant changes in flux (Fig s6D).

Untargeted compound fluxes in patient breath showed no separation between normal, benign or cancer groups using unsupervised PCA (Fig s6B). Splitting patients by age as before also revealed no separation by PCA in untargeted data (Fig s7D) but very clear separation along component 1 of PLS-DA (Fig s7E). This separation was driven by compounds with retention time of 11.1 (and 15.7 mins with variable importance projection (VIP) scores over 2 (Fig s7F).

### Breath volatile flux identifies cancer and benign tumours in age-controlled groups

Once patients were controlled for age (all patients grouped in ≤50 and >50 groups) this resulted in uneven distribution of patients across groups, with patients in the cancer group skewed to >50. Patient numbers were as follows: >50 = Normal, 8; benign, 8; Cancer, 17, and ≤50 = Normal, 12; benign, 13; Cancer, 2 (Fig s6C).

Within the >50 group and using only targeted compounds, chloroform and styrene fluxes were significantly different (Fig 3G &H, respectively). Chloroform fluxes were increased relative to normal patients in the >50s for benign and cancer patients (Fig 3G). All >50s in the normal/healthy group actively metabolised, or consumed, chloroform whereas in the benign and cancerous groups there was, on average, metabolic production, or release in the breath (Fig 3G). Metabolic production of styrene was significantly increased in the benign >50s group compared to both cancer and normal groups (Fig 3H). No other compounds were observed to be significantly different between groups in the >50s (Fig s8A-K).

In the ≤50 patient group, useful comparisons were only possible for benign vs normal patients due to only 2 patients with cancer (table 1). Butanone was significantly increased in benign patients ≤50 compared with normal patients (Fig s9I), no other compounds were shown to be significantly different between health statuses (Fig s9A-K).

Untargeted analysis was performed on the >50s patient groups. Unsupervised PCA revealed little separation among groups (Fig s10A) but clear separation was achieved with supervised PLS-DA; benign patients were clearly separated from normal patients with cancer patients between each group along component 1 (Fig 3I). The top 15 compounds based on VIP scores from PLS-DA (Fig s10B) and random forest analysis (Fig s10C) were tested for significance and only a compound with retention time 9.3 and tentative ID of isopropyl alcohol (0.940 MoNA match, see table 2) was found to be significant (Fig 3J), suggesting over fitting of the PLS-DA (Fig 3I).

**Table 2.** Tentative identification of significantly altered compounds.

| Retention time | CAS | Tentative ID | MoNA match |
| --- | --- | --- | --- |
| 9.3 | 67-63-0 | Isopropyl Alcohol | 0.94 |
| 35.06 | 5780-36-9 | 2-(methylthio)thiophene | 0.709 |

Further untargeted analysis to investigate any potential compounds capable of separating benign and normal patients revealed retention times of 19.7, 15.7 and 21.73 mins as important using PLS-DA (Fig s10B), however these compounds were not present in >50% of patients.

Untargeted analysis comparing normal vs benign revealed clear separation of groups along component 1 (Fig s10D). A compound at 35.06 mins with m/z 109 was significantly increased in benign patients vs normal patients (Fig s10F) but no other compounds were significantly changed in the flux values. Comparing benign vs cancer also revealed clear separation along component 1 using PLS-DA (Fig s10G) with VIP score presented in Fig s10H, with no flux values identified as significant.

Overall, once age-based differences were addressed, chloroform, styrene, and isopropyl alcohol allowed differentiation between normal, benign, and cancer patients. We found no relationship between BMI and VOC fluxes and, because of the small group sample sizes, it was not possible to determine any significant effects of comorbidity or impacts of drugs such as statins upon breath flux profiles.

### VOC flux separated cancer grade

Breast cancer grades are given by pathologists to aid in prognosis and treatment. Grades 1-3 represent increasingly invasive and aggressive profiles with grade 1 being most like normal tissue and grade 3 being most abnormal [21]. We were interested in whether breath VOCs could distinguish differences in grade and so we examined volatile fluxes relative to the grades assigned to each patient by the Magnolia Centre breast clinic staff. Due to collection methodology, we were unable to recruit patients based on grade and therefore had 3 grade 1 patients, 9 grade 2 and 6 grade 3 (table 1). No grade information was collected for 2 patients (Identified as VOC 25 and 57). For VOC 25 tumour size was small (4mm) preventing grading but was included in grade 1 as it was consistent with this group based on VOC profiles. VOC 57 had multiple tumours (table 1) and was not included in any group. VOC 39 presented complex pathology with multiple tumours and TNM staging not provided (table 1) and so, while categorised in grade 2, it presented as an outlier and was excluded from analysis.

Grades were not clearly separated using PCA analysis (Fig s11A) however, supervised PLS-DA revealed separation of grade 1 from grade 2 and 3 along component 1 (Figure 4A). Volatile flux analysis revealed 2 significantly different compounds between groups; chloroform was significantly increased in grade 2 vs grade 1 (Fig 4B) and DMS was decreased in both grade 2 and 3 compared with grade 1 (Fig 4C). Both chloroform and DMS had the highest VIP scores (over 1.5) from the PLS-DA (Fig s11B).

**Figure 4.**
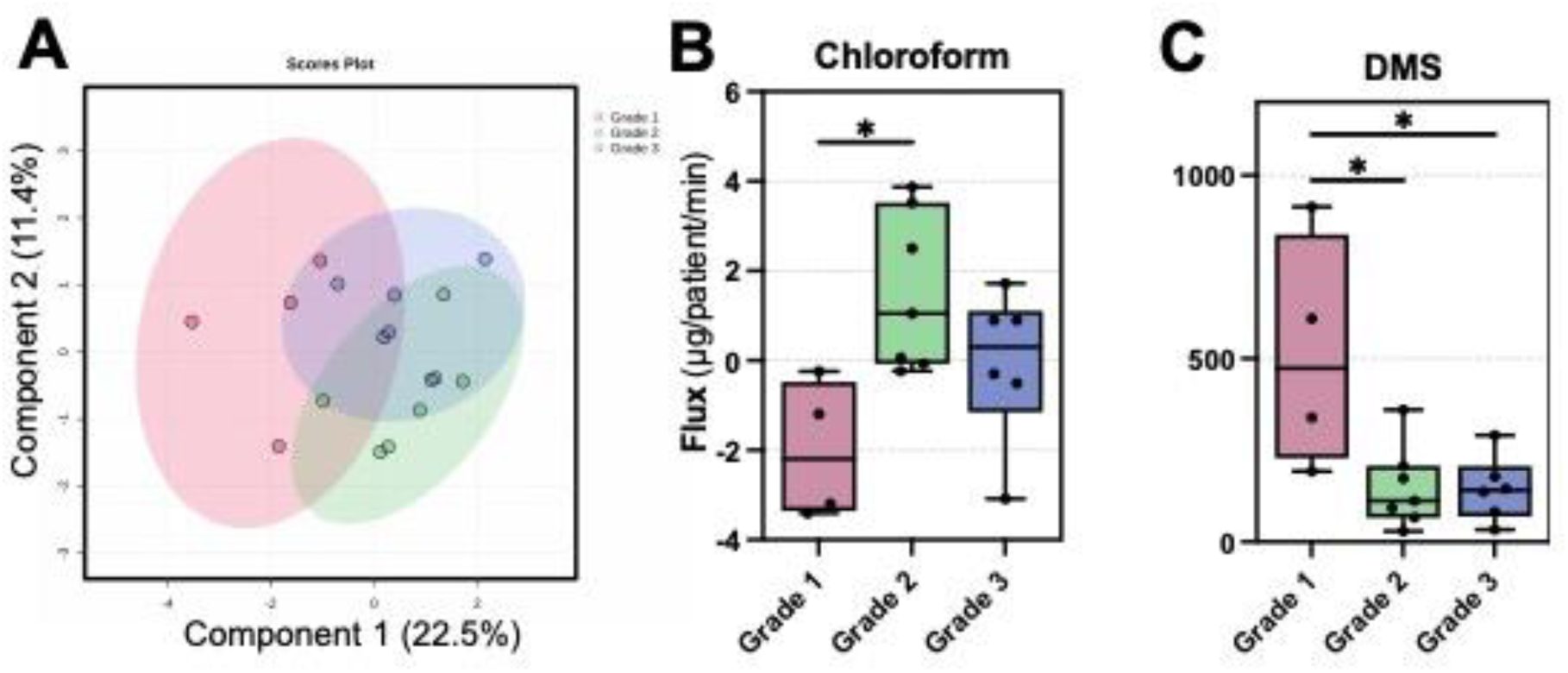
Breath volatile flux can separate cancer grades and groups when controlling for age. **(A)** Partial least squared discriminant analysis (PLS-DA) of over 50s cancer patients seperated by cancer grade select volatile flux. Boxplots of chloroform **(B)** and dimethyl sulfide (DMS) (C) flux values in µg/patient/min. Boxplots show median ± Tukey distribution (n= 20, 21, 19 for normal, benign and cancer groups respectively). One-way AN OVA with Tukey post hoc analysis was performed; *p = <0.05.

Untargeted analysis on these same grades revealed no separation using PCA (Fig s11C) but very clear separation of all groups using PLS-DA mostly along component 1 (Fig s11D). Compound VIP scores from component 1 are shown in Fig s11E, with the primary driver being high levels of a compound with retention time of 16.1 in stage 1 cancers, which was identified as DCM, supporting targeted analysis. Compounds with retention times of 28.38 (primary mass of 77, closest match: phenyl ethyl acetate, 0.736 in MoNA) and 33.37 (primary mass of 105, with no close matches) were both top drivers of separation in the PLS-DA but these compounds were not significantly changed in the raw flux values, with the majority of samples showing no presence (no compound detected).

### Breath volatile flux identifies cancer and benign tumours in age-controlled groups

Combining all biomarkers from targeted/untargeted analyses which were identified as either i) significantly altered in the flux values or ii) defining of patient health status we applied multivariate exploratory receiver operator curve (ROC) analysis using monte-carlo cross validation to generate an optimal panel of biomarkers for ROC analysis. 13 compounds were compared for patients in the >50 group, including the targeted panel, isopropyl alcohol and the compound with retention time of 35 mins (m/z109) with a tentative ID of 2-(methylthio)thiophene (table 2).

Comparison of normal vs cancer patients revealed top compounds by multivariate exploratory ROC as chloroform, acetone, isopropyl alcohol, butanone and styrene (Fig s12A). Using these 5 compounds ROC analysis generated and area under the curve (AUC) of 0.934 (with 95% confidence interval (CI) of 0.721-1) when comparing these two groups (Fig 5A).

**Figure 5.**
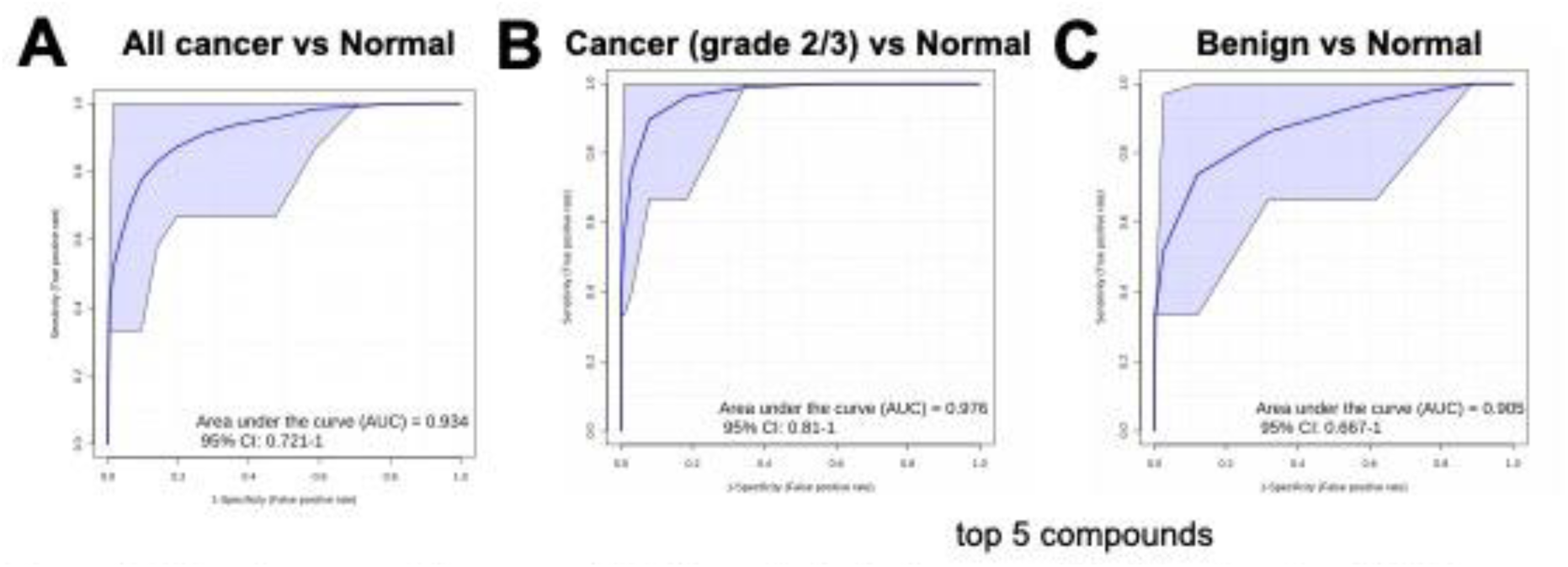
Receiver operator curve (ROC) analysis in the over 50s. Random forest ROC curves generated using the top 5 compounds identified using multivariate exploratory ROC analysis. Top 5 compounds were identified based on average importance score from random forest Monte-Carlo cross validation. **(A)** All patients compared with normal patients (area under the curve, AUC = 0.934). **(B)** Cancer patients with a grade 2 or 3 tumour compared with normal patients (AUC = 0.976) and **(C)** benign patients vs normal patients (AUC = 0.905).

We determined that grade 1 breast cancer patients were significantly different from grade 2/3 (Figure 4B & C) and therefore pooled grade 2 and 3 patients to compare accuracy in diagnosing these patients from normal patients in the > 50s. Multivariate exploratory ROC identified chloroform, acetone, isopropyl alcohol, DMS and butanone as the top 5 compounds (Fig s12B). ROC analysis with these 5 compounds generated and AUC of 0.976 with 95% CI of 0.81-1 (Fig 5B).

Multivariate exploratory ROC analysis of benign vs normal flux revealed Styrene, the compound at 35mins, acetone, chloroform and butanone as the top 5 compounds (Fig s12C). ROC analysis with these 5 compounds generated and AUC of 0.905 with 95% CI of 0.667-1 (Fig 5C).

## Discussion

Breath volatile flux, incorporating concurrent measurement of inhaled and exhaled breath, has revealed several important findings in breast cancer detection and classification. Our results demonstrate that accounting for age-related metabolic variations is crucial for accurate interpretation of breath volatile signatures. Furthermore, our findings demonstrate that our dual-sampling approach, combined with age stratification, enables more precise identification of disease-specific metabolic signatures than traditional single-point breath sampling methods.

We have reviewed a range of cancer-specific breath research and shown that compound classes, in particular, aldehydes, alkanes and aromatics, can define breast cancer [5]. In this current study, styrene was shown to have utility as a biomarker in identifying benign tumours (Fig 3H, s12C) and cancer (Fig s12A). However, other than styrene, we report no aldehydes or alkanes as descriptors of cancer or benign disease.

In the previous review we presented a compound-specific breath VOC cancer panel which informed the biomarkers targeted in this research. Butanone was predicted as a key positive biomarker of cancer in this compound-specific analysis and has been shown in this study to be a key indicative of cancer status in those >50 yrs (butanone is related to aldehydes via the carbonyl group, but the attachment position is quite different) [5]. Butanone has been reported previously as a biomarker of breast cancer in urine [22] however, this is the first report of this compound in the breath of breast cancer patients. Butanone has however been reported in the breath of lung cancer patients [5] and has been shown to be more abundant in later stage lung cancer [23].

To our knowledge this is the first reported example of chloroform being identified as a key VOC, predictive of cancer, released in the breath. The ability to identify this compound as having diagnostic capabilities is only made possible through our flux approach (Fig 1) since variability in the environment is substantial. The underlying basis for chloroform metabolism in patients with tumours (benign or cancerous) is not yet understood however we have observed modulation by breast cancer cells of chlorinated VOCs (MeCl and chloroform) under conditions of hypoxia [7] and chemotherapeutic stress [8]. Previously, we have reported chloroform significantly increased in ageing HEK-293t, glial cells and in DJ-1 *drosophila* mutants [19].

Isopropyl alcohol has been reported as a compound distinctive of cancer in-vivo [15], observed in the urine of breast cancer patients [22] and breath [24], lung cancer and gastric cancer [25], suggesting shared metabolic pathways across malignancies. Isopropyl alcohol is also observed in the breath of lung cancer patients [5] and could be linked to ketone metabolism [26], elevated levels in both butanone and isopropyl alcohol could therefore be indicative of similar metabolic responses. Isopropyl alcohol could however be an exogenous compound, commonly found in hospitals as a cleaning agent and the utility of this biomarker would need to be assessed.

The age-related variations in metabolic patterns, particularly in chloroform flux, represent a novel finding in breath analysis. This observation highlights the importance of age-stratification in breath-based diagnostic approaches and a control which we have not seen implemented previously in breast cancer breath research [5, 27–31]. This finding will not only improve diagnostic specificity but also lays the groundwork for future studies to incorporate demographic stratifications as standard practice in breath analysis. Our previously observed modulation of chloroform in models of ageing [19] further implicates this VOC as important in age-related processes.

Particularly notable is our ability to distinguish between cancer grades using breath volatile fluxes, a capability not previously reported in breast cancer breath analysis. This finding suggests potential applications in non-invasive monitoring of disease progression and establishes a direct link between pathophysiology and breath volatile profiles.

The methodological advantages of our flux-based approach address several key challenges recently identified by Bajo-Fernández et al. (2024) in their comprehensive review of GC-MS-based breath metabolomics [6]. While traditional approaches struggle with environmental contamination and sampling standardization, our dual-sampling method effectively controls for these variables, enhancing the reliability of biomarker identification. The impact of compound retention within the body due to variable compound partial coefficients may confound the utility of individual biomarkers, however, these would need to be assessed in a case by case basis.

Our findings demonstrate relevance when compared to recent developments in point-of-care breath testing. Phillips et al. (2024) recently reported a rapid breath test for breast cancer screening [20]; while their approach prioritizes immediate results, our flux-based methodology offers potentially superior diagnostic accuracy through comprehensive metabolic profiling. This suggests complementary roles for different breath analysis approaches in clinical settings.

## Conclusion

This study presents several significant advances in breath-based cancer diagnostics. Our novel flux-based approach, incorporating simultaneous measurement of inhaled and exhaled breath, has revealed age-dependent metabolic signatures and cancer-specific volatile markers. The identification of chloroform as an age-influenced cancer indicator, alongside the discovery of grade-specific volatile patterns, represents a substantial step forward in non-invasive cancer diagnostics. The ability to distinguish between cancer grades using breath volatiles offers promising applications for disease monitoring and treatment response assessment. These findings, combined with our robust methodology for controlling environmental variables, establish a foundation for more reliable breath-based diagnostic tools. Future research should focus on validating these biomarkers in larger cohorts and investigating their potential role in longitudinal patient monitoring. Our results demonstrate that breath volatile flux analysis, when properly controlled for age and environmental factors, can provide valuable insights into cancer metabolism and potentially serve as a complementary diagnostic tool in clinical settings.

## Supporting information

Complete_figures

## Data Availability

All data produced in the present study are available upon reasonable request to the authors

## Acknowledgements

The authors would like to thank the Elsie May Sykes award from the York and Scarborough NHS trust which funded this work. The work was also supported by the White Rose Mechanistic Biology Doctoral Training Program, supported by the Biotechnology and Biological Science Research Council (BBSRC) BB/M011151/1. The authors would like to acknowledge the support provided by Mark Bentley in the University of York Department of Biology workshop.

## Conflict of interest

The authors declare that the research was conducted in the absence of any commercial or financial relationships that could be construed as a potential conflict of interest.

## Notes

### Competing Interest Statement

The authors have declared no competing interest.

### Funding Statement

This study was funded by Elsie May Sykes NHS fund

### Author Declarations

Ethics committee of York and Scarborough NHS trust gave ethical approval for this work Ethics committee of University of York gave ethical approval for this work

