## Supplementary material for "Breath Volatile Flux Reveals Age-Dependent Metabolic Markers for Breast Cancer Detection": Complete_figures

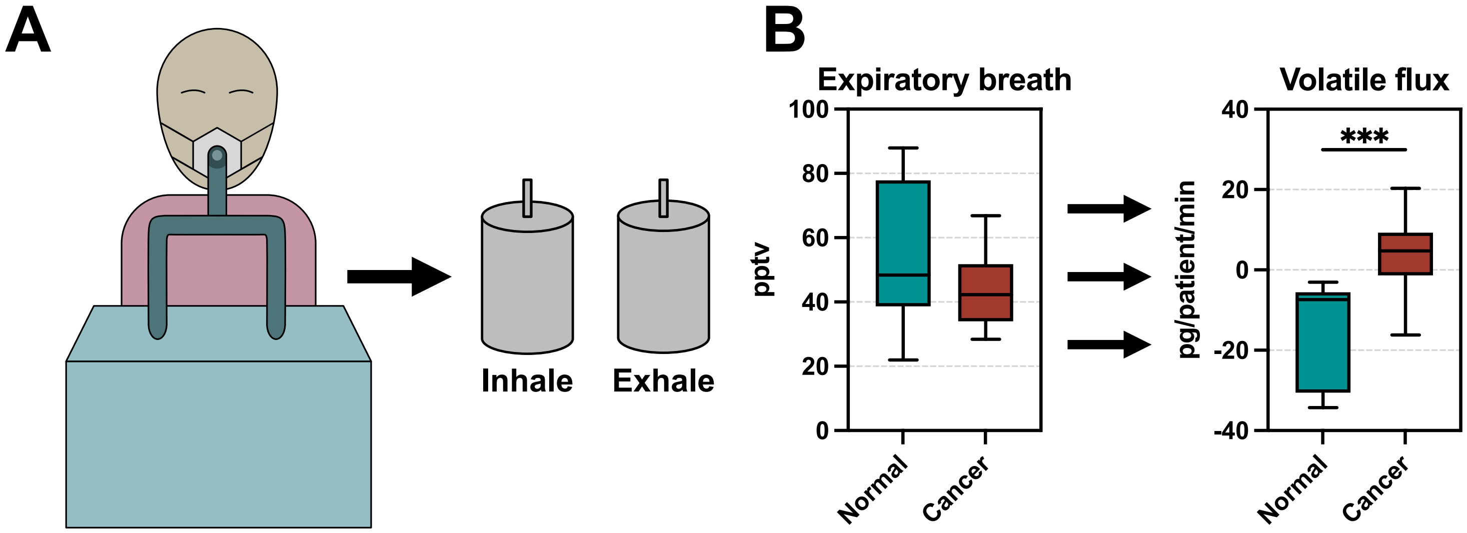


Figure 1. **Overview of breath volatile flux. (A)** Breath is captured over 5 mins of tidal breathing at rest. Air is drawn into the circuit through a one way valve, driven by patients breathing a T-valve. Two samples are generated, an inhale and exhale sample. **(B)** An example of a single volatile compound (chloroform) in parts per trillion volume in the exhaled breath of normal patients and breast cancer patients with grade 2 or 3 tumours vs the volatile flux of the same compound in pg/patient/min. Unpaired t-test, n= 8 for normal patients and n=14 for cancer patients; *** = $p<0.001$.

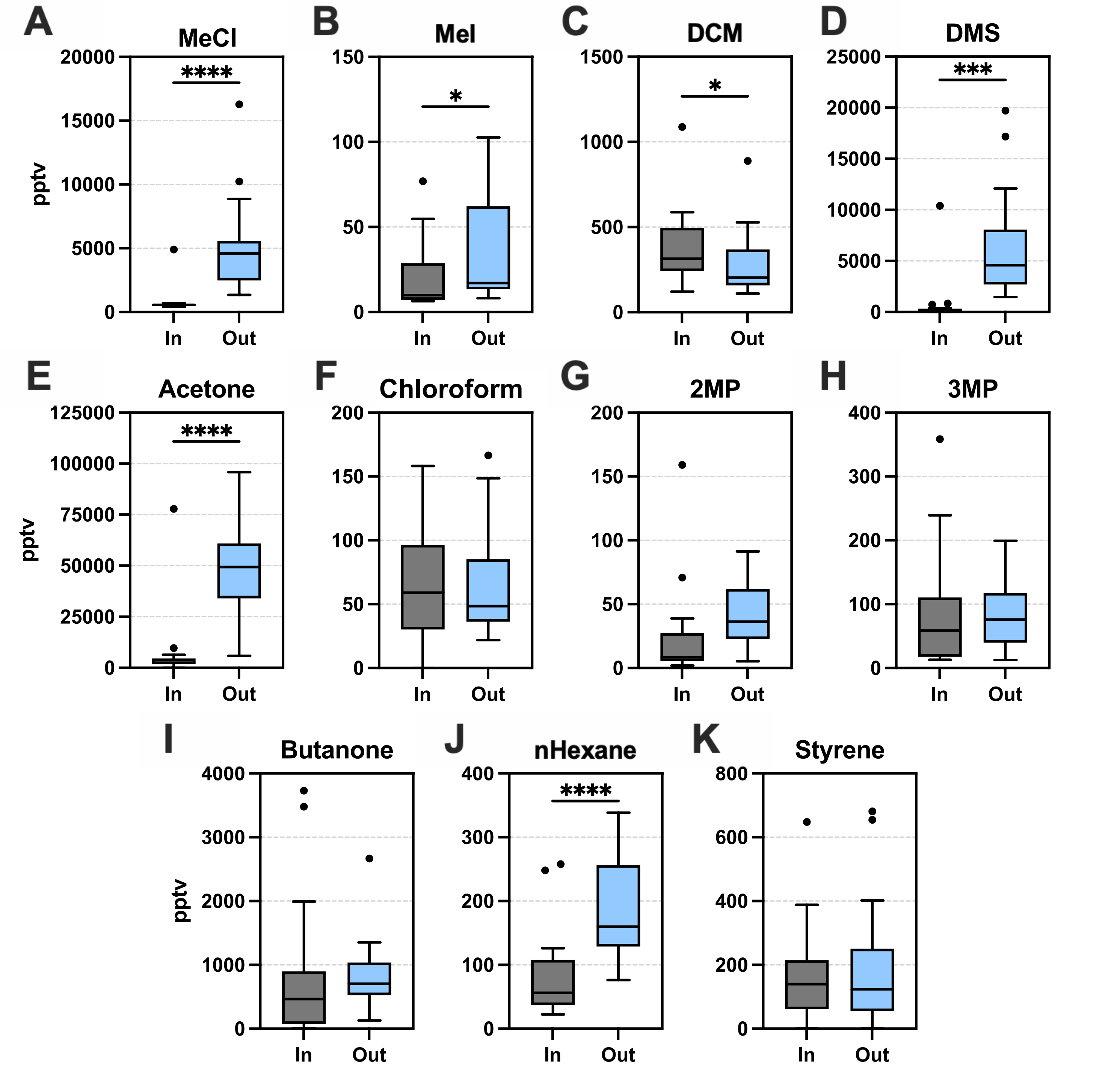


Supplementary Figure 2. **Targeted compound concentrations in Inhaled (In) versus exhaled (Out) breath.** Boxplots for 20 female patients from the 'normal' group, mixed ages. Boxplots show median ± Tukey distribution, in and out breath sample are collected at the same time for each patient. Paired t-test was performed; ****p = <0.0001, ***p = <0.001, *p = <0.05.

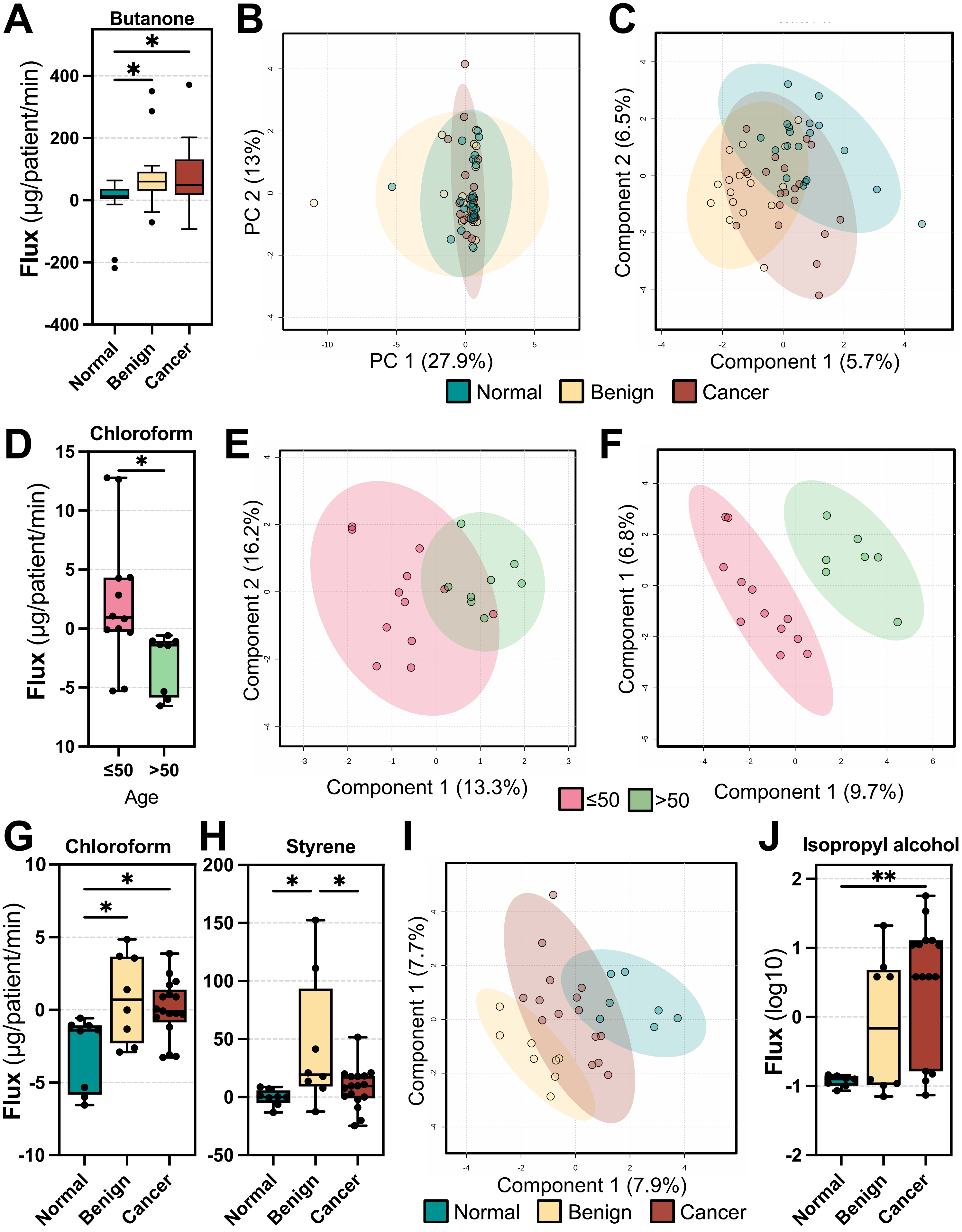


Figure 3. **Volatile flux in the breath of normal, benign and breast cancer patients. (A)** Butanone flux is significantly increased in benign and cancer patients vs normal patients in µg/patient/min. **(B)** Principle component analysis and **(C)** partial least squared discriminant analysis (PLS-DA) of 12 select volatile compound flux values. **(D)** Boxplot of normal group chloroform flux values in µg/patient/min for 50 and under and over 50s. **(E)** PLS-DA for 12 select volatile compounds and **(F)** untargeted analysis for normal group based on age, 50 and under vs over 50 for select volatile flux. **(G)** Chloroform and **(H)** styrene flux in µg/patient/min in the over 50s. **(I)** PLS-DA of untargeted analysis in the over 50s. **(J)** Isopropyl alchohol flux (log10) in the over 50s. Boxplots show median ± Tukey distribution (n= 20, 21, 19 for normal, benign and cancer groups respectively). Unpaired t-test was performed for **D**; **p = < 0.01, *p = <0.05.

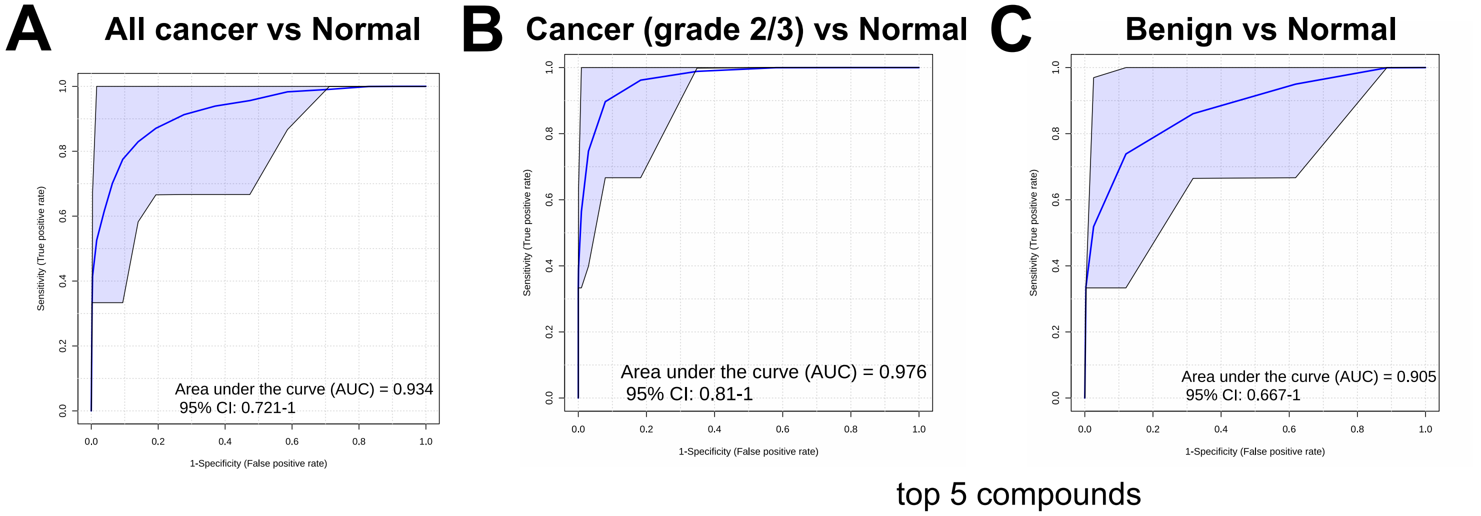


Figure 5. **Receiver operator curve (ROC) analysis in the over 50s.** Random forest ROC curves generated using the top 5 compounds identified using multivariate exploratory ROC analysis. Top 5 compounds were identified based on average importance score from random forest Monte-Carlo cross validation. **(A)** All patients compared with normal patients (area under the curve, AUC = 0.934), **(B)** Cancer patients with a grade 2 or 3 tumour compared with normal patients (AUC = 0.976) and **(C)** benign patients vs normal patients (AUC = 0.905).

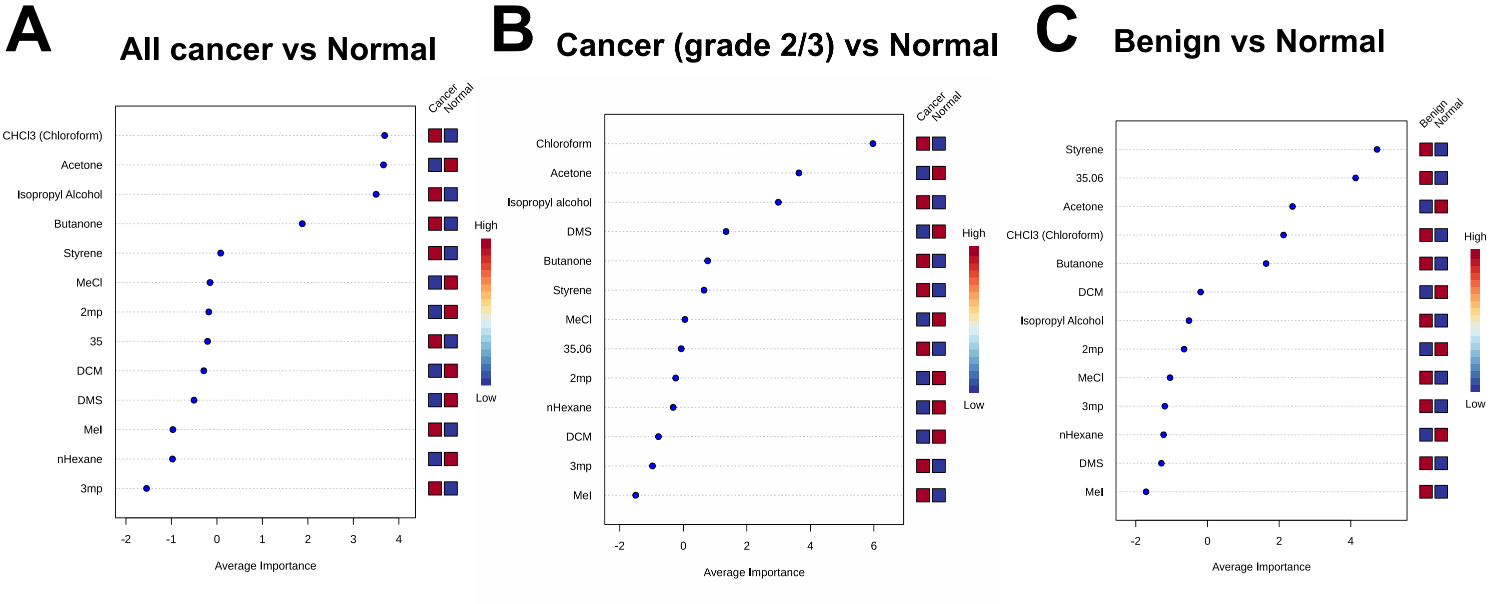


Supplementary Figure 12. **Multivariate exploratory receiver operator curve (ROC) analysis.** Random forest ROC curves generated using Monte-Carlo cross validation and top compounds used to build ROC curves presented in Figure 5. **(A)**

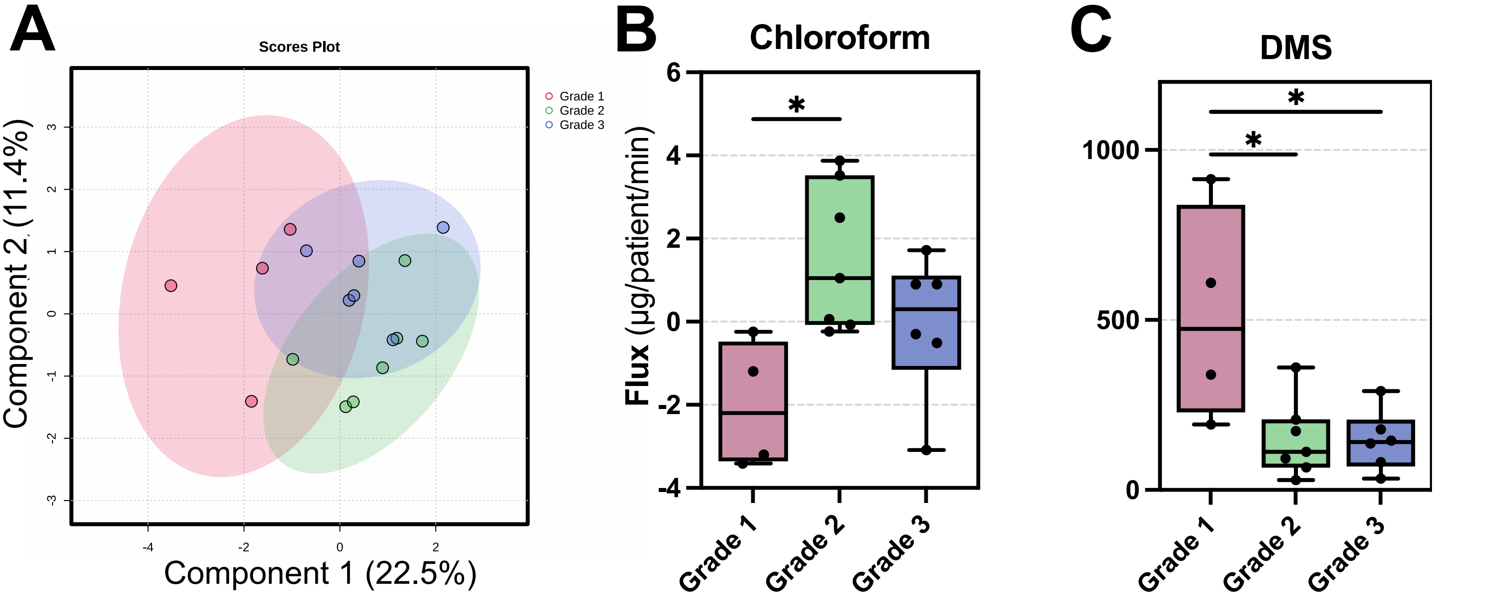


Figure 4. **Breath volatile flux can separate cancer grades and groups when controlling for age (A)** Partial least squared discriminant analysis (PLS-DA) of over 50s cancer patients seperated by cancer grade select volatile flux. Boxplots of chloroform **(B)** and dimethyl sulfide (DMS) **(C)** flux values in µg/patient/min. Boxplots show median ± Tukey distribution (n= 20, 21, 19 for normal, benign and cancer groups respectively). One-way ANOVA with Tukey post hoc analysis was performed; *p = <0.05 .

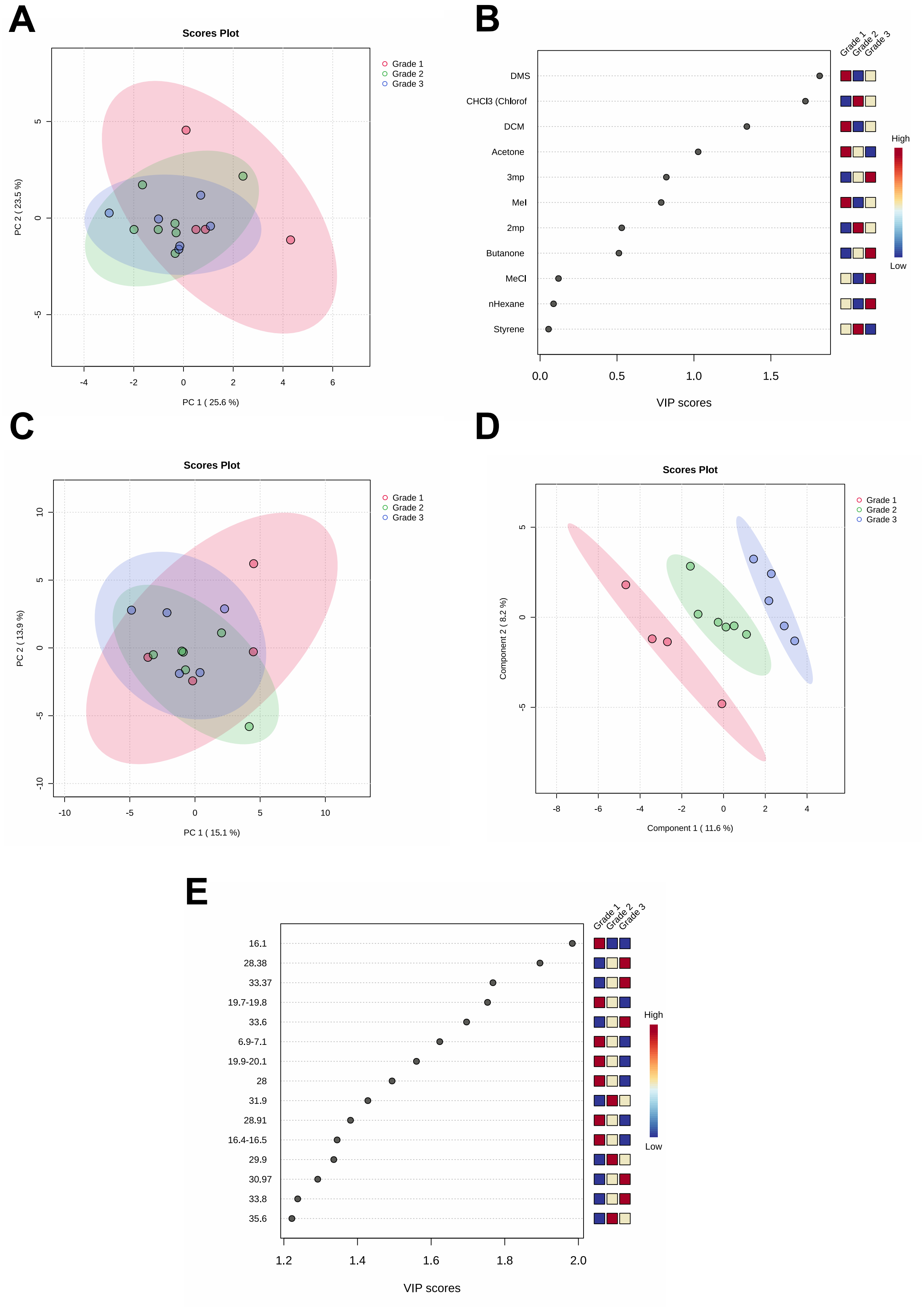


Supplementary Figure 11. **Volatile flux based on cancer grade.** Breath volatile flux in the over 50s split by cancer grade and multivariate analysis. **(A)** PCA of targeted compounds and associated VIP scores for targeted analysis **(B)**. **(C)** PCA for untargeted flux analysis. **(D)** PLS-DA for untargeted flux analysis **(E)** PLS-DA VIP scores from untargeted flux analysis

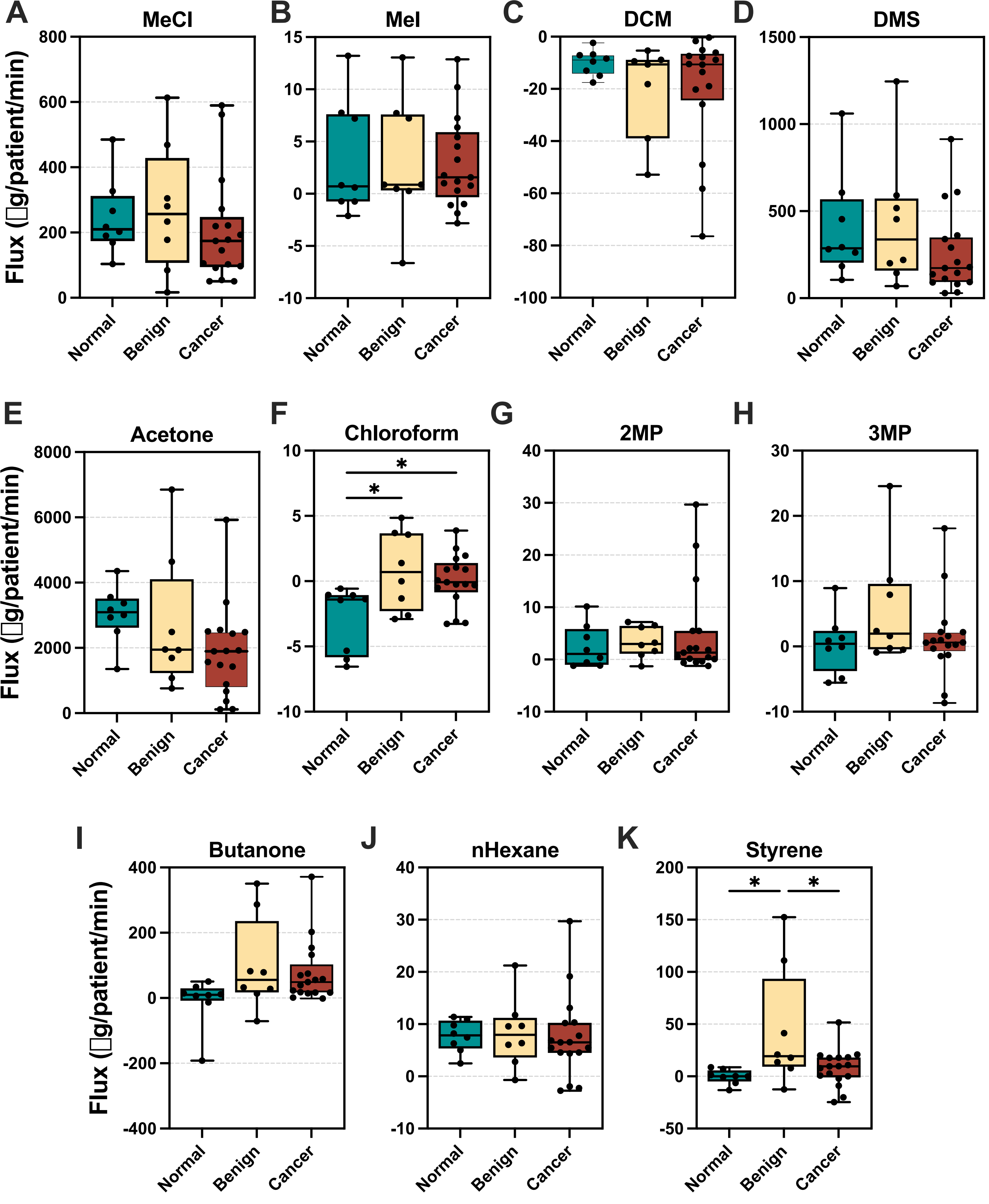


Supplementary Figure 8. **Volatile flux in over 50s for select compounds.** Boxplots for female patients from all groups, age 51 and over (n= 8 normal, 8 benign, 17 cancer) Boxplots show median ± Tukey distribution, in and out breath sample are collected at the same time for each patient. One way ANOVA with Tukey post hoc test performed; ****p = <0.0001, ***p = <0.001, **p = <0.01; *p = <0.05.

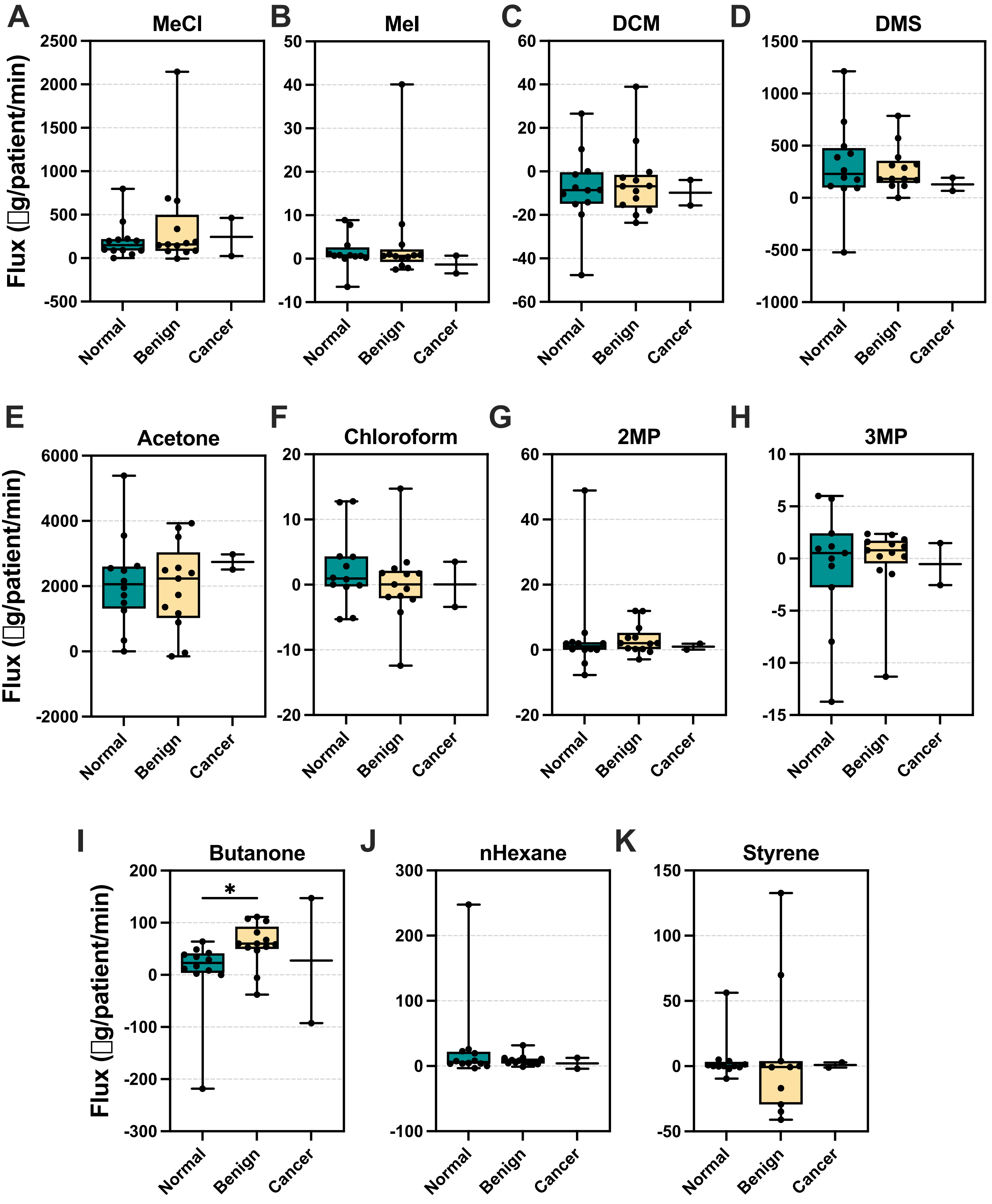


Supplementary Figure 9. **Volatile flux in 50 and under for select compounds.** Boxplots for female patients from all groups, age 50 and under (n= 12 normal, 13 benign, 2 cancer) Boxplots show median ± Tukey distribution, in and out breath sample are collected at the same time for each patient. One way ANOVA with Tukey post hoc test performed; ****p = <0.0001, ***p = <0.001, **p = <0.01; *p = <0.05.

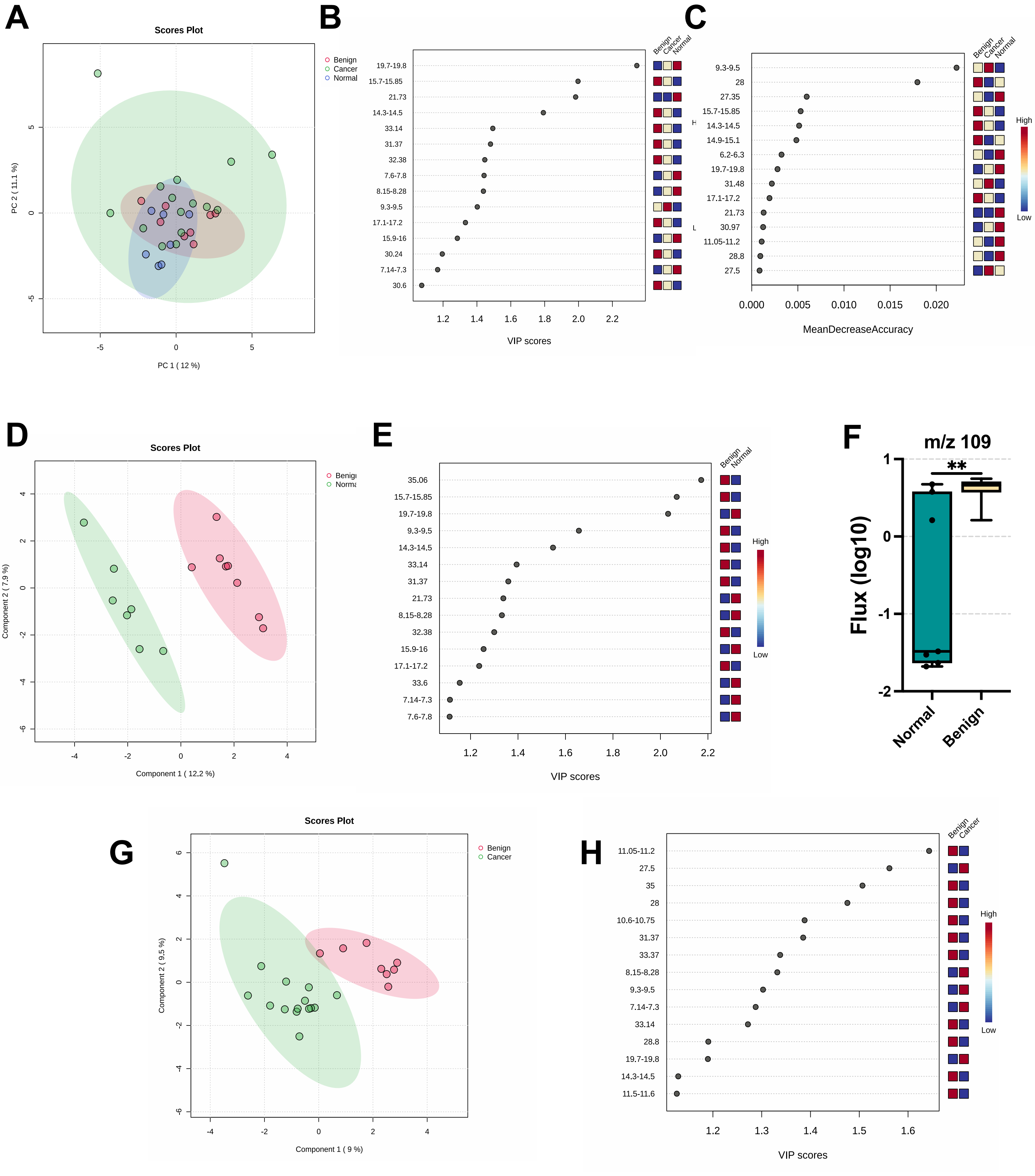


Supplementary Figure 10. **Untargeted analysis of breath flux in over 50s. (A)** Principle component anaysis of untargeted compound flux in the over 50s for normal (n=8), benign (n= 8) and cancer (n=15). **(B)** VIP scores from PLS-DA for untargeted analysis in the over 50s as in A. **(C)** Mean decrease accuracy scores from untargeted analysis using random forest analysis. **(D)** PLS-DA of beign vs normal patients with associated VIP scores **(E)**. **(F)** Normalised flux (log 10) of a compounds with mass 109 and retention time 35.06. **(G)** PLS-DA of benign vs cancer and associated VIP scores **(H)**. Students t-test performed for **(F),** **p= <0.001.

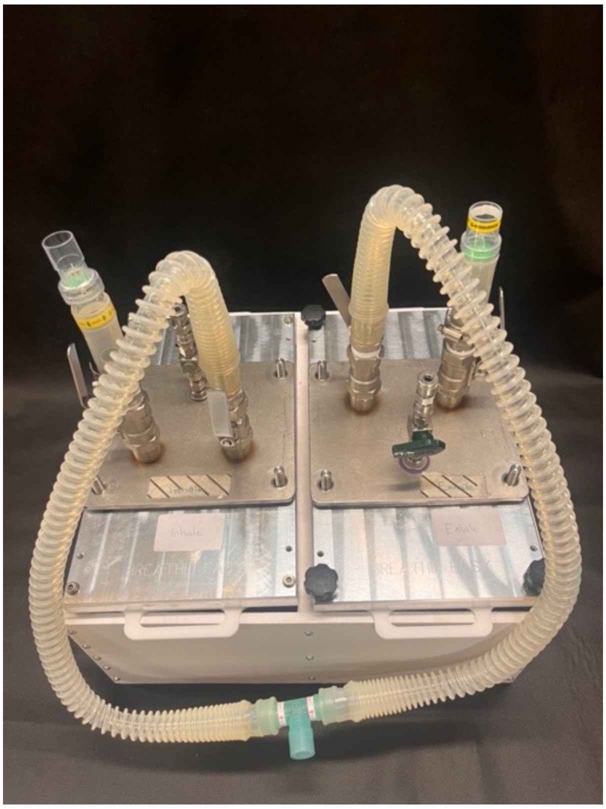


Supplementary Figure 1. **Picture of breath flux prototype device.**

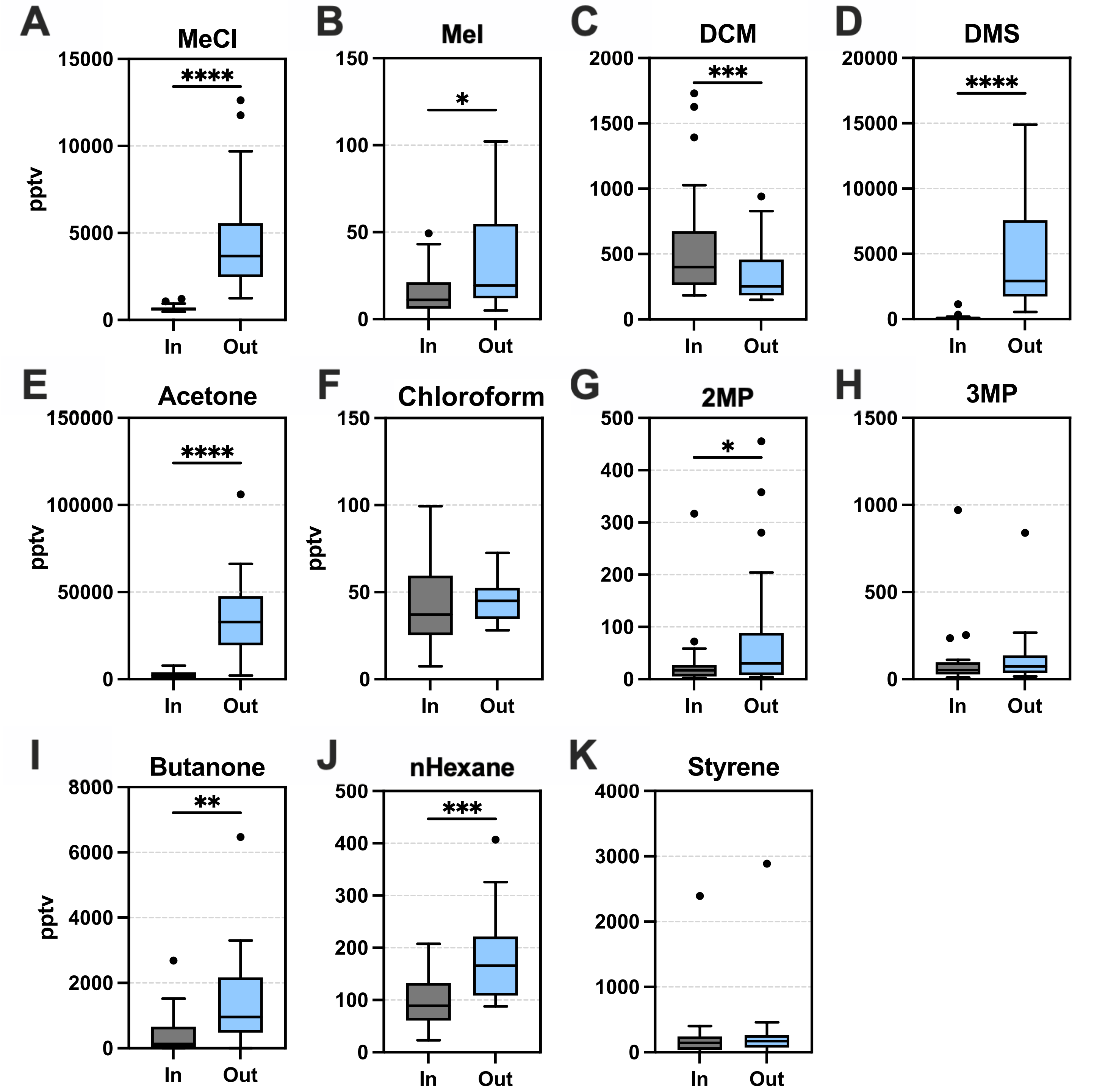


Supplementary Figure 3. **Targeted compound concentrations in Inhaled (In) versus exhaled (Out) breath.** Boxplots for 19 female patients from the 'cancer' group, mixed ages. Boxplots show median ± Tukey distribution, in and out breath sample are collected at the same time for each patient. Paired t-test was performed; ****p = <0.0001, ***p = <0.001, **p = <0.01; *p = <0.05.

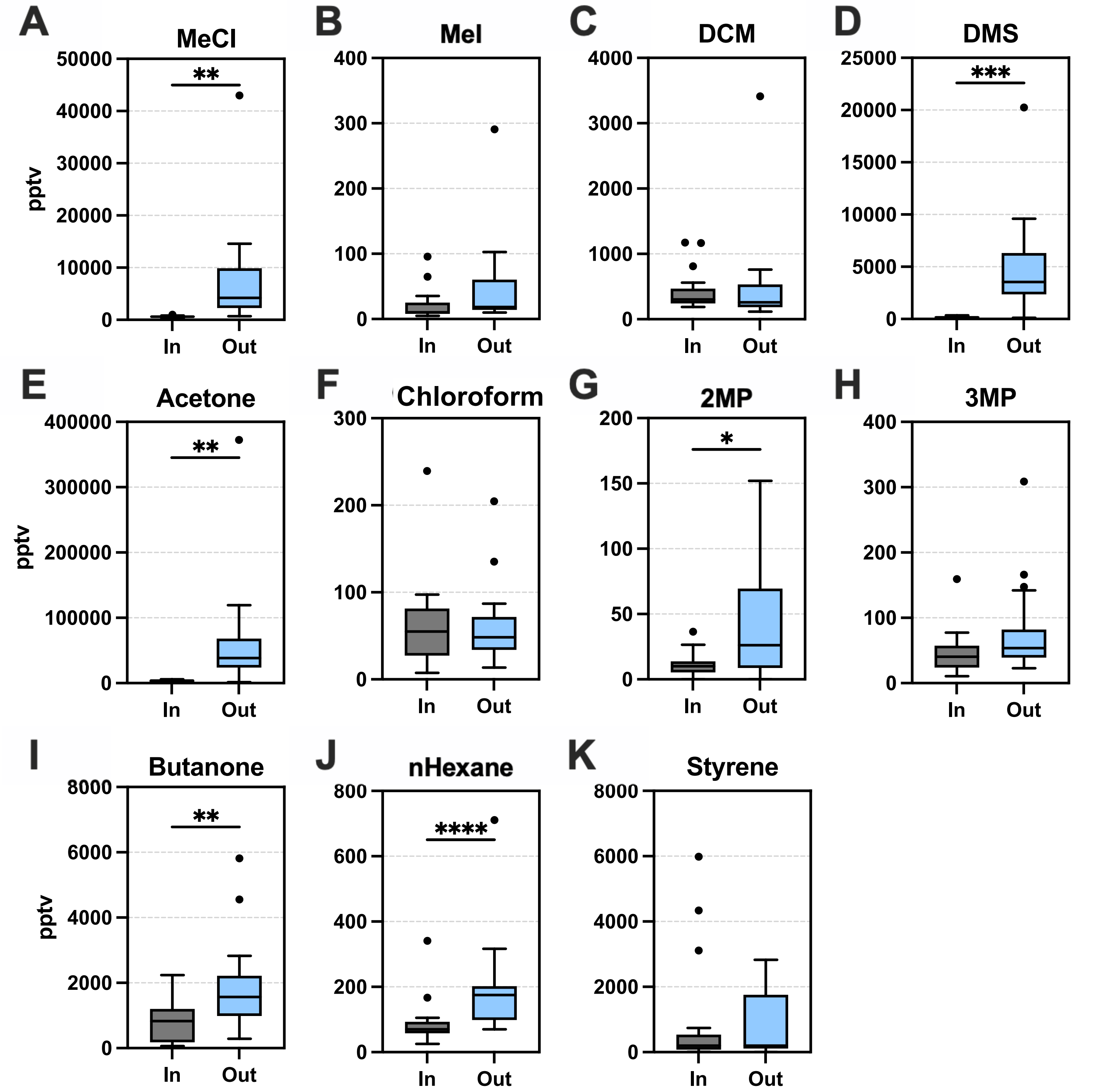


Supplementary Figure 4. **Targeted compound concentrations in Inhaled (In) versus exhaled (Out) breath.** Boxplots for 21 female patients from the 'benign' group, mixed ages. Boxplots show median ± Tukey distribution, in and out breath sample are collected at the same time for each patient. Paired t-test was performed; ****p = <0.0001, ***p = <0.001, **p = <0.01; *p = <0.05.

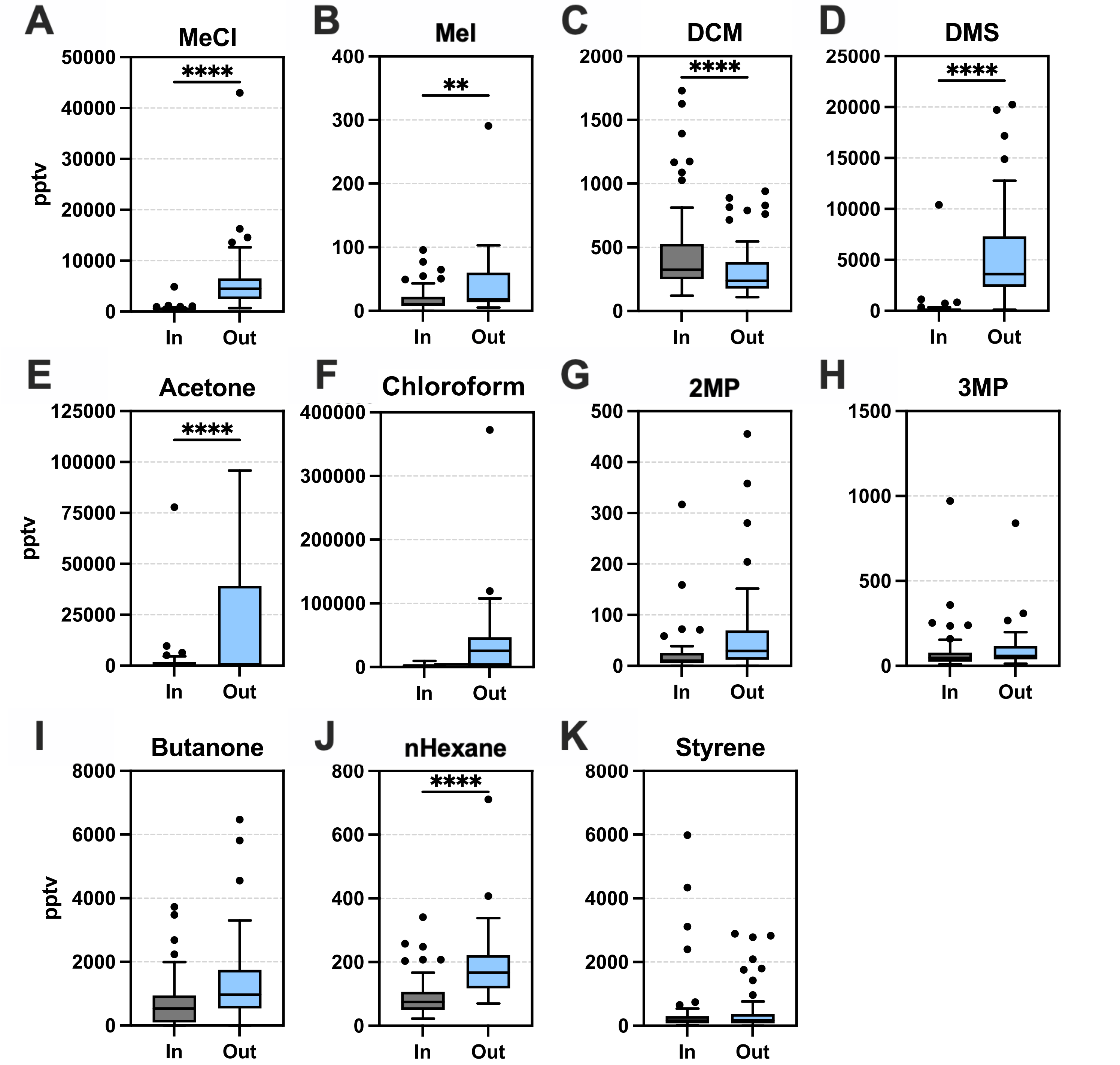


Figure 2. **Targeted compound concentrations in Inhaled (In) versus exhaled (Out) breath.** Boxplots for 60 female patients, mixed ages (19-94 years). Boxplots show median ± Tukey distribution, in and out breath sample are collected at the same time for each patient. Paired t-test was performed; ****p = <0.0001, **p = 0.01.

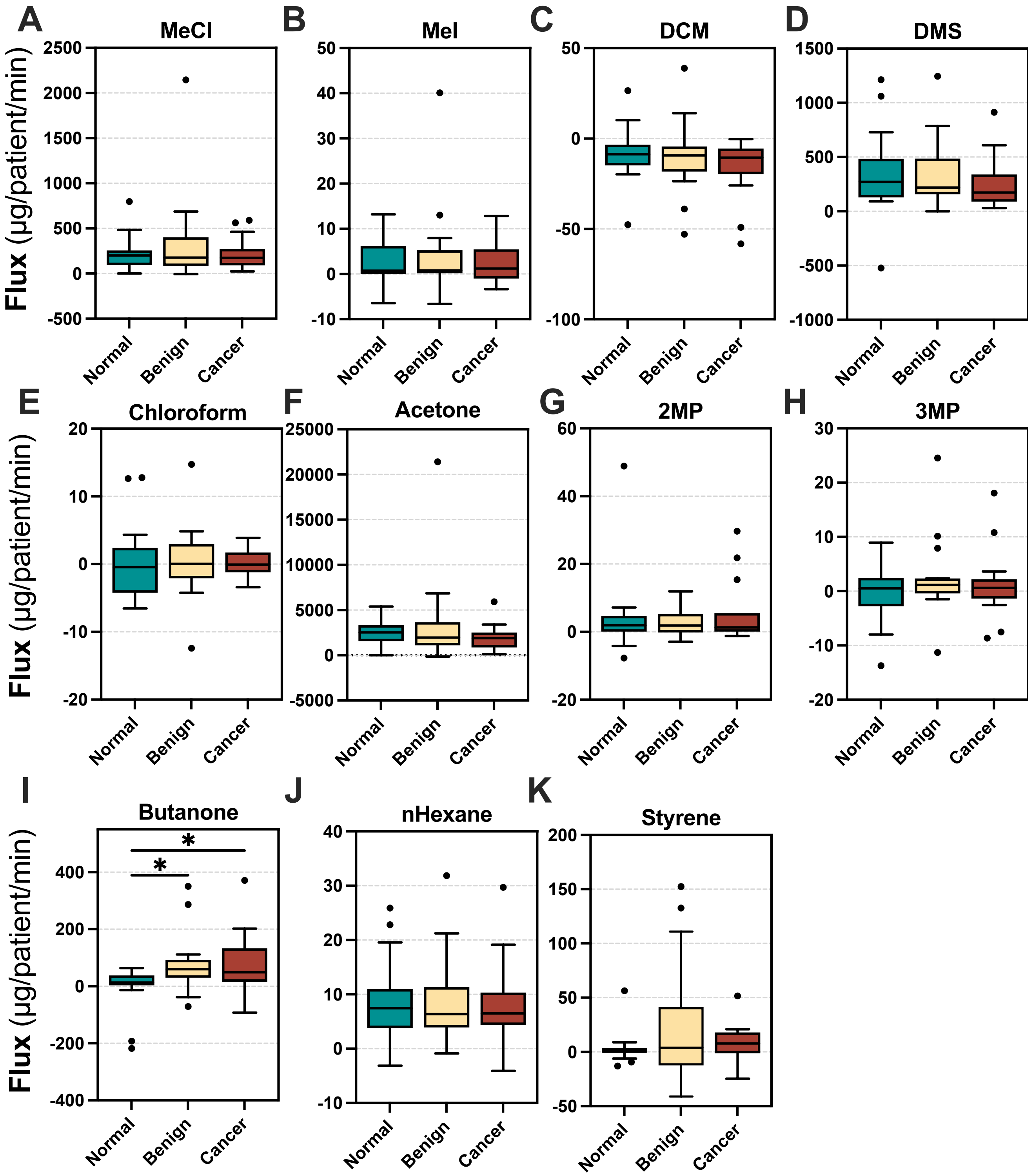


Supplementary Figure 5. **Breath volatile flux for normal, benign and breast cancer patients.** Boxplots for 60 female patients from the all groups (n= 20 normal, 21 benign, 19 cancer), mixed ages (33-94 years). Boxplots show median ± Tukey distribution, in and out breath sample are collected at the same time for each patient. One way ANOVA with Tukey post hoc test performed; ****p = <0.0001, ***p = <0.001, **p = <0.01; *p = <0.05.

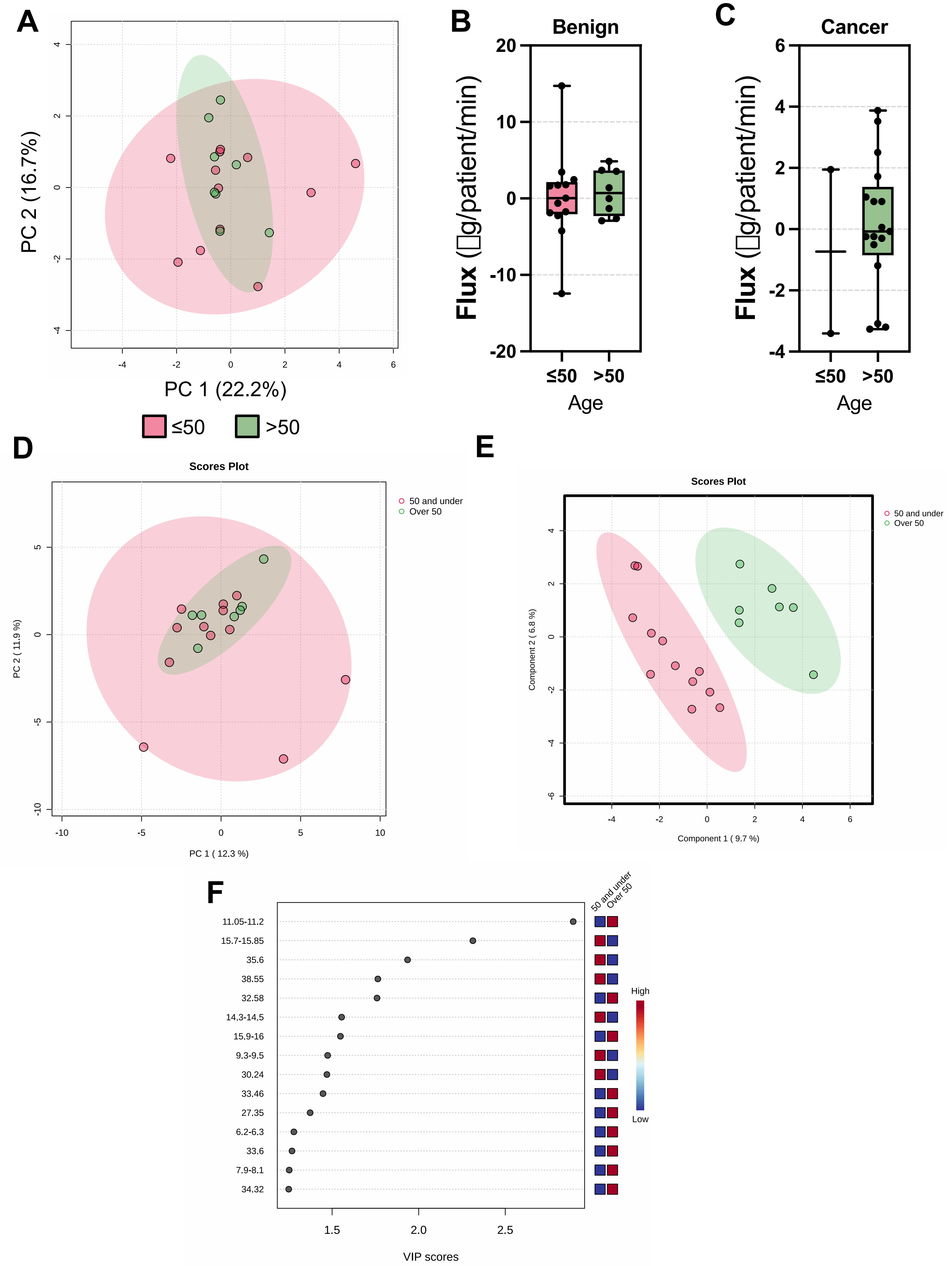


Supplementary Figure 7. **Volatile flux in the breath of normal, benign and breast cancer patients based on age. (A)** Principle component analysis (PCA) of normal patients in 50 and under ages vs over 50s (n=20). **(B)** chloroform flux in benign group based on age (n=21) and **(C)** chloroform flux for cancer group based on age (n=19). **(D)** PCA of compounds identified in untargeted analysis in normal patients based on age. **(E)** partial least squared analysis of untargeted data from **(D)**. **(F)** variable importance projection (VIP) scores for the top 15 compounds for PLS-DA analysis in **(E)**.

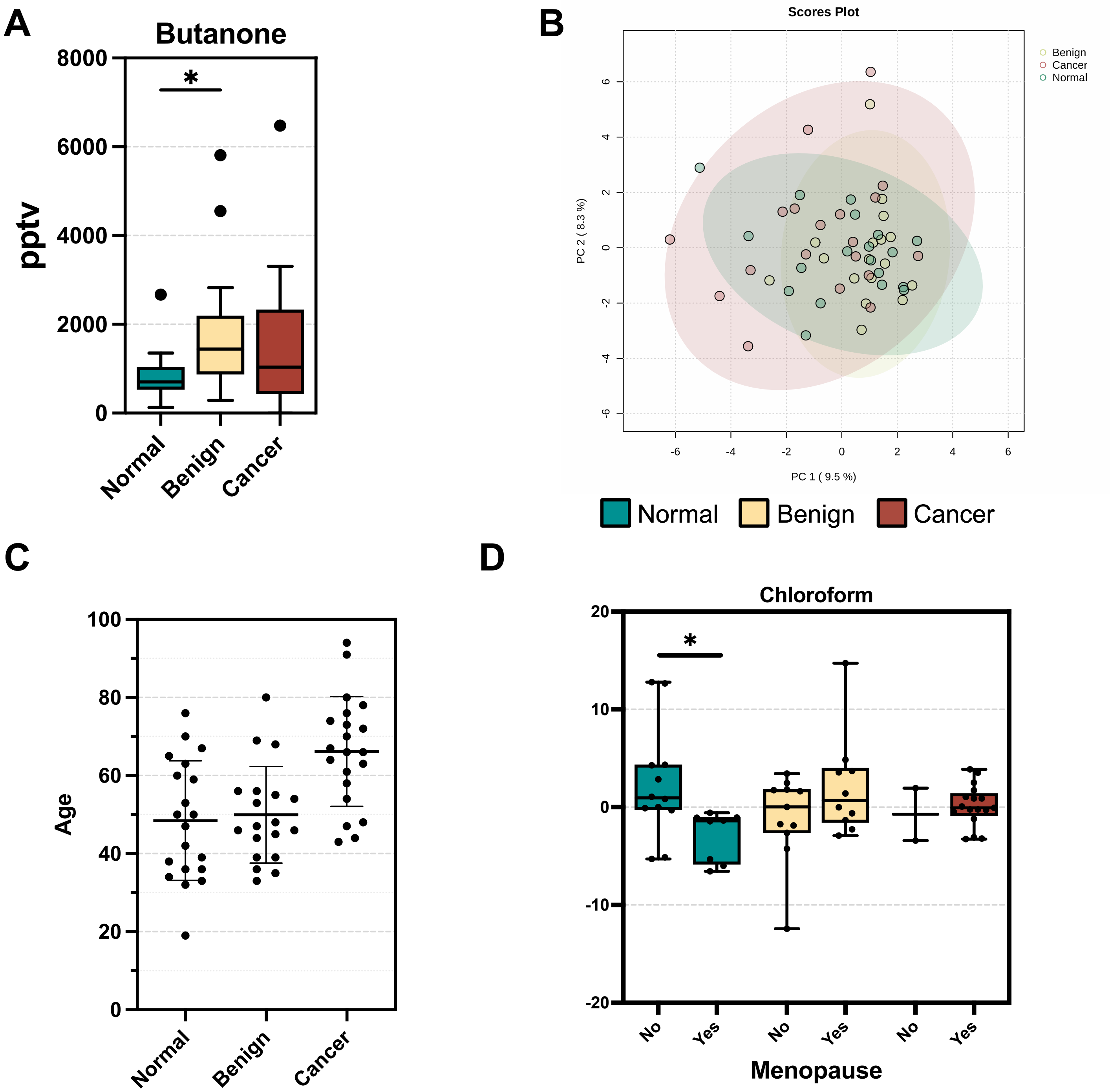


Supplementary Figure 6. **Volatile flux in the breath of normal, benign and breast cancer patients. (A)** Chloroform in parts per trillion volume (pptv) in the exhaled breath of patients (normal, n=20; benign, n=21; cancer, n=19). **(B)** Principle component analysis (PCA) of volatile flux from compounds identified through untargeted analysis. **(C)** Age (years) distribution of patients within each group. **(D)** Chloroform flux in µg/patient/min for patients within normal, benign or cancer group dependent upon menopause status. One-way ANOVA with tukey post hoc analysis was performed for **A.** *p = <0.05.
